# Precision Gestational Diabetes Treatment: Systematic review and Meta-analyses

**DOI:** 10.1101/2023.04.15.23288459

**Authors:** Jamie L Benham, Véronique Gingras, Niamh-Maire McLennan, Jasper Most, Jennifer M Yamamoto, Catherine E Aiken, Susan E Ozanne, RM Reynolds ADA/EASD PMDI

## Abstract

We hypothesized that a precision medicine approach could be a tool for risk-stratification of women to streamline successful GDM management. With the relatively short timeframe available to treat GDM, commencing effective therapy earlier, with more rapid normalization of hyperglycaemia, could have benefits for both mother and fetus. We conducted two systematic reviews, to identify precision markers that may predict effective lifestyle and pharmacological interventions. There were a paucity of studies examining precision lifestyle-based interventions for GDM highlighting the pressing need for further research in this area. We found a number of precision markers identified from routine clinical measures that may enable earlier identification of those requiring escalation of pharmacological therapy. Whether there are other sensitive markers that could be identified using more complex individual-level data, such as ‘omics’, and if these can be implemented in clinical practice remains unknown. These will be important to consider in future studies.

## INTRODUCTION

Gestational diabetes (GDM) is the most common pregnancy complication, occurring in 3% to 25% of pregnancies globally^1^. GDM is associated with significant short- and long-term risks to both mothers and babies, including adverse perinatal outcomes, future obesity, type 2 diabetes and cardiovascular disease^1-3^. The landmark Australian Carbohydrate Intolerance Study in Pregnant Women (ACHOIS) demonstrated that effective treatment of GDM reduces serious perinatal morbidity^4^.

Current treatment guidelines for management of GDM assume homogeneous treatment requirements and responses, despite the known heterogeneity of GDM aetiology^5-8^. Standard care includes diet and lifestyle advice at a multi-disciplinary clinic, home blood glucose monitoring at least four times per day, clinic reviews every two to four weeks, and then progression to pharmacological treatment with metformin, glyburide and/or insulin if glucose targets are not met. Around a third of women cannot maintain euglycaemia with lifestyle measures alone and require treatment escalation to a pharmacological agent^3^. Yet current treatment pathways often take 4-8 weeks to achieve glucose targets. This delay resulting in continued exposure to hyperglycaemia poses a significant risk of accelerated fetal growth^9,10^. Previous research has suggested that maternal characteristics including body mass index (BMI) ≥30 kg/m^2^, family history of type 2 diabetes, prior history of GDM and higher glycated haemoglobin (HbA1c) increase the likelihood of need for insulin treatment in GDM^11^, indicating the potential for risk-stratification of women to streamline successful GDM management. There is emerging evidence that precision biomarkers predict treatment response in type 2 diabetes, which has similar heterogeneity to GDM^12,13^ and thus gives rationale to investigate whether a similar precision approach could be successful in optimizing outcomes in GDM.

To address this knowledge gap, we conducted two systematic reviews of the available evidence for precision markers of GDM treatment. We aimed to determine (i) which precision diet and lifestyle interventions delivered in addition to standard of care enable achievement of glucose targets with lifestyle measures alone, (ii) which patient-level characteristics or factors predict whether glucose targets can be achieved in women treated with diet and lifestyle alone, and in women receiving oral agents for treatment of GDM.

The Precision Medicine in Diabetes Initiative (PMDI) was established in 2018 by the American Diabetes Association (ADA) in partnership with the European Association for the Study of Diabetes (EASD). The ADA/EASD PMDI includes global thought leaders in precision diabetes medicine who are working to address the burgeoning need for better diabetes prevention and care through precision medicine^14^. This systematic review is written on behalf of the ADA/EASD PMDI as part of a comprehensive evidence evaluation in support of the 2nd International Consensus Report on Precision Diabetes Medicine^15^.

## METHODS

The systematic reviews and meta-analyses were performed as outlined *a priori* in the registered protocols (PROSPERO registration IDs CRD42022299288 and CRD42022299402). The Preferred Reporting Items for Systematic reviews and Meta-Analyses (PRISMA) guidelines^16^ were followed. Ethical approval was not required as these were secondary studies using published data.

### Literature Searches, Search Strategies and Eligibility Criteria

Search strategies for both reviews were developed based on relevant keywords in partnership with scientific librarians (see Supplementary Text S1 for full search strategies). We searched two databases (MEDLINE and EMBASE) for studies published from inception until January 1^st^, 2022. We also scanned the references of included manuscripts for inclusion as well as relevant reviews and meta-analyses published within the past two years for additional citations.

For both systematic reviews we included studies (randomized or non-randomized trials, and observational studies) published in English and including women ≥16 years old with diagnosed GDM, as defined by the study authors. For the first systematic review (precision diet and lifestyle interventions), we included studies with any behavioural intervention (e.g., exercise, diet, motivational interviewing) over and above standard care compared to a control group receiving standard care only. For the second systematic review (precision predictors of need for pharmacological interventions to achieve glucose targets), we included studies using pharmacological therapy to treat GDM (e.g., insulin, metformin, sulphonylurea) compared to a control group receiving standard care with diet and lifestyle measures, or taking oral agents before progression to insulin. For both reviews, we included any relevant reported outcomes; maternal (e.g., treatment adherence, hypertensive disorders of pregnancy, gestational weight gain, mode of birth), neonatal (e.g., birthweight, macrosomia, shoulder dystocia, preterm birth, neonatal hypoglycaemia, neonatal death), cost efficiency or acceptability. We excluded studies with a total sample size <50 participants to ensure sufficient data to interpret the effect of precision markers. We also excluded studies published before or during 2004, in order to consider studies with standard care similar to ACHOIS^4^.

### Study selection and data extraction

The results of our two searches were imported separately into Covidence software (Veritas Health Innovation, Australia, available at www.covidence.org) and duplicates were removed. Two reviewers independently reviewed identified studies. First, they screened titles and abstracts of all references identified from the initial search. In a second step, the full-text articles of potentially relevant publications were scrutinized in detail and inclusion criteria were applied to select eligible articles. Reason for exclusion at the full text review stage was documented. Disagreement between reviewers was resolved through consensus by discussion with the group of authors.

Two reviewers independently extracted relevant information from each eligible study, using a pre-specified standardized extraction form. Any disagreement between reviewers was resolved as outlined above.

Data extracted included first author name, year of publication, country, study design, type and details of the intervention when applicable, number of cases/controls or cohort groups, total number of participants and diagnostic criteria used for GDM. Extracted data elements also included outcomes measures, size of the association (Odds Ratio (OR), Relative Risk (RR) or Hazard Ratio (HR)) with corresponding 95% Confidence Interval (CI) and factors adjusted for, confounding factors taken into consideration and methods used to control covariates. We prioritized adjusted values where both raw and adjusted data were available. Details of precision markers (mean (standard deviation) for continuous variables or N (%) for categorical variables) including BMI (pre-pregnancy or during pregnancy), ethnicity, age, smoking status, comorbidities, parity, glycaemic variables (e.g., oral glucose tolerance test (OGTT) diagnostic values, HbA1c), timing of GDM diagnosis, history of diabetes or of GDM, and season were also extracted.

### Quality assessment (risk of bias and GRADE assessments)

We first assessed the quality and risk of bias of each individual study using the Joanna Briggs Institute (JBI) critical appraisal tools^17^. A Grading of Recommendations, Assessment, Development, and Evaluations (GRADE) approach was then used to review the total evidence for each precision marker, and the quality of the included studies to assign a GRADE certainty to this body of evidence (high, moderate, low and/or very low)^18^. Quality assessment was performed in duplicate and conflicts were resolved through consensus.

### Statistical analysis

Where possible, meta-analyses were conducted using random effects models for each precision marker available. The pooled effect size (mean difference for continuous outcomes and ORs for categorical outcomes) with the corresponding 95% CI were computed. The heterogeneity of the studies was quantified using I^2^ statistics, where I^2^ >50% represents moderate and I^2^ >75% represents substantial heterogeneity across studies. Publication bias was assessed with visual assessment of funnel plots. Statistical analyses were performed using Review Manager software [RevMan, Version 5.4.1, The Cochrane Collaboration, Copenhagen, Denmark].

## RESULTS

### Study selection and study characteristics

PRISMA flow charts (Figures 1A and 1B) summarize both searches and study selection processes.

**Figure 1.**
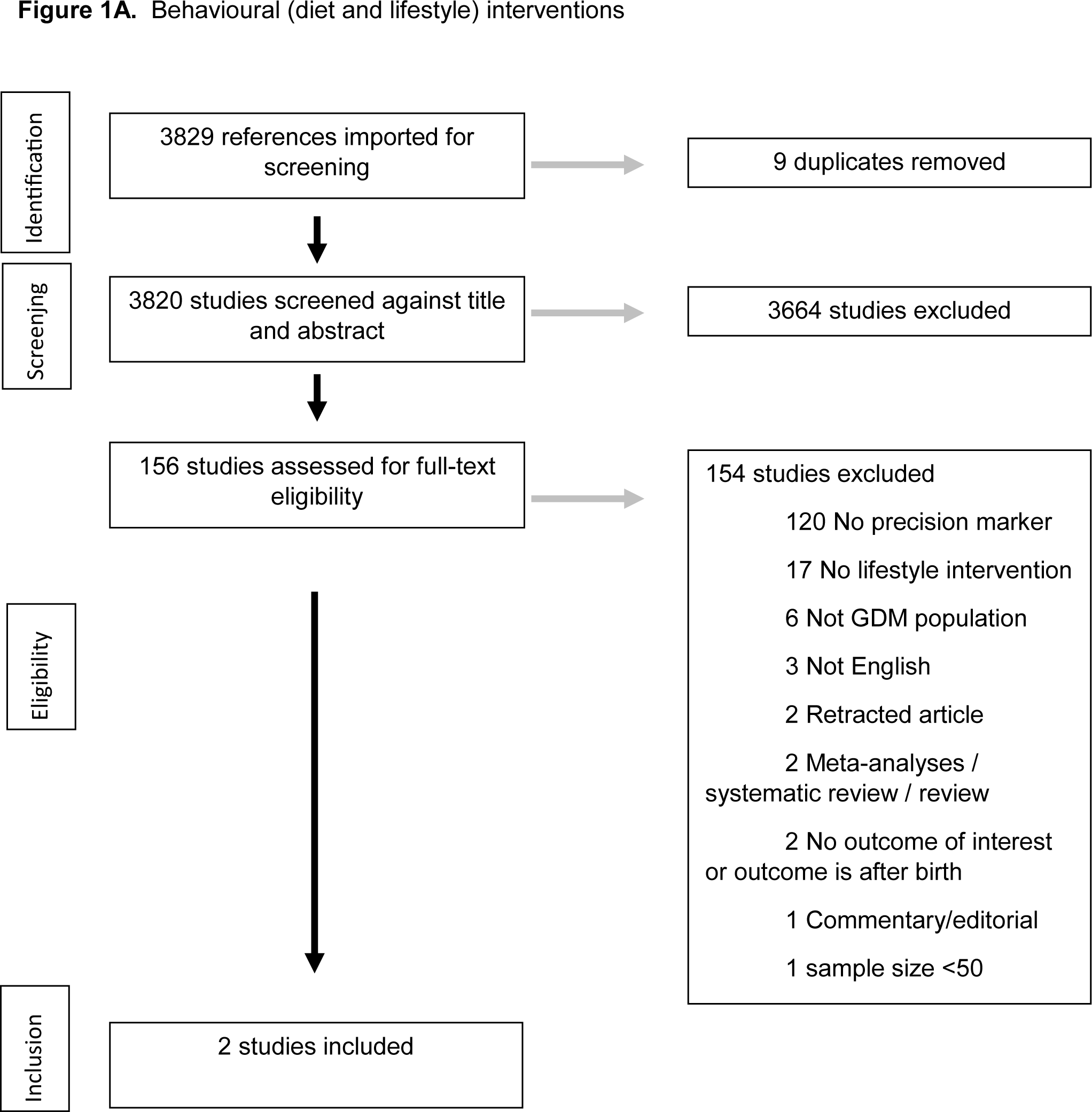
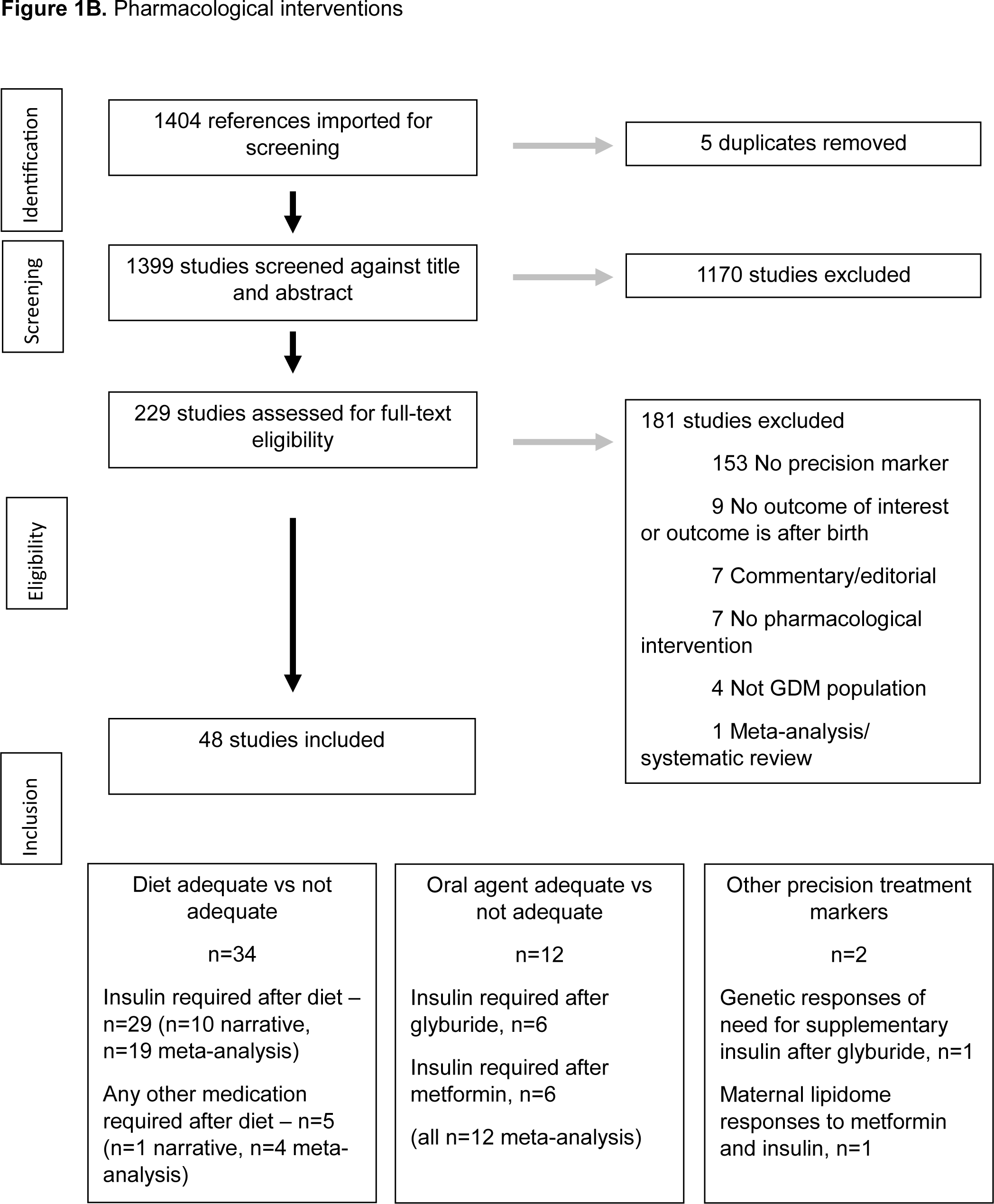
Preferred Reporting Items for Systematic Reviews and Meta-Analyses (PRISMA) flow diagrams for the two systematics reviews: **A)** behavioural (diet and lifestyle) interventions and **B)** pharmacological interventions.

For the first systematic review (precision diet and lifestyle interventions), we identified 2 eligible studies (n=2,354 participants), which were randomized trials from USA and Singapore (Table 1A)^19,20^.

**Table 1A.**
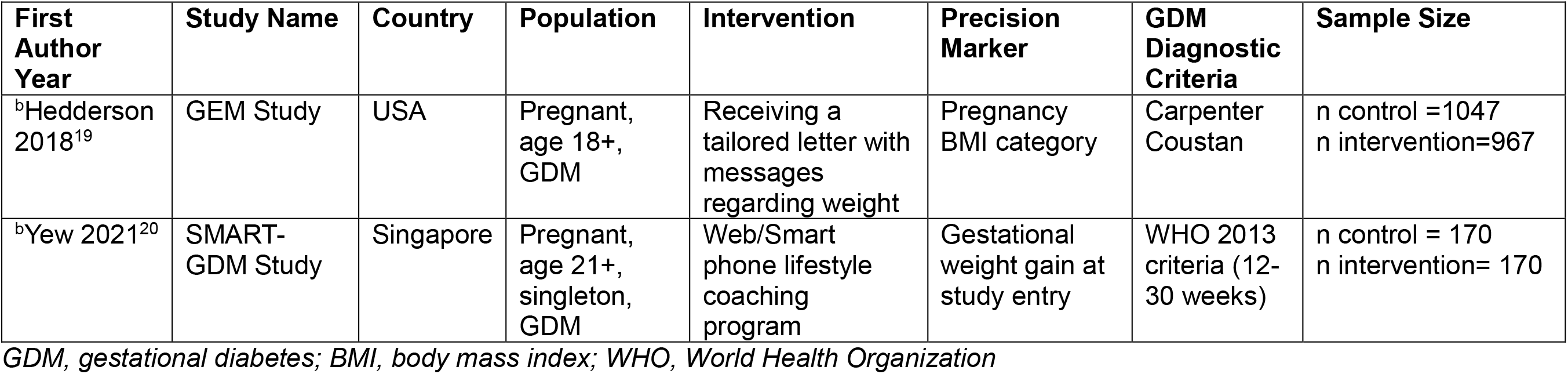
Summary of included studies in the two systematic reviews. Precision behavioural (diet and lifestyle) interventions

| First Author Year | Study Name | Country | Population | Intervention | Precision Marker | GDM Diagnostic Criteria | Sample Size |
| --- | --- | --- | --- | --- | --- | --- | --- |
| <sup>b</sup>Hedderson 2018<sup>19</sup> | GEM Study | USA | Pregnant, age 18+, GDM | Receiving a tailored letter with messages regarding weight | Pregnancy BMI category | Carpenter Coustan | n control =1047<br>n intervention=967 |
| <sup>b</sup>Yew 2021<sup>20</sup> | SMART-GDM Study | Singapore | Pregnant, age 21+, singleton, GDM | Web/Smart phone lifestyle coaching program | Gestational weight gain at study entry | WHO 2013 criteria (12-30 weeks) | n control = 170<br>n intervention= 170 |

*GDM, gestational diabetes; BMI, body mass index; WHO, World Health Organization*

For the second systematic review (precision predictors of need for pharmacological interventions to achieve target glucose levels), we identified 48 eligible studies (n=25,724 participants) (Table 1B)^21-68^.

**Table 1B.**
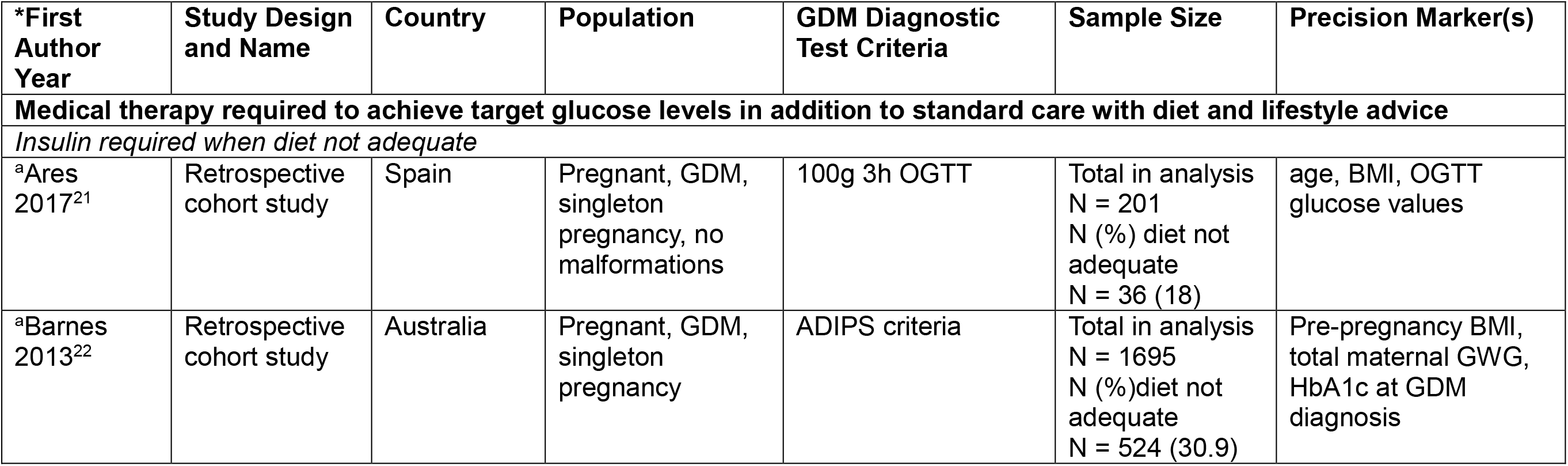
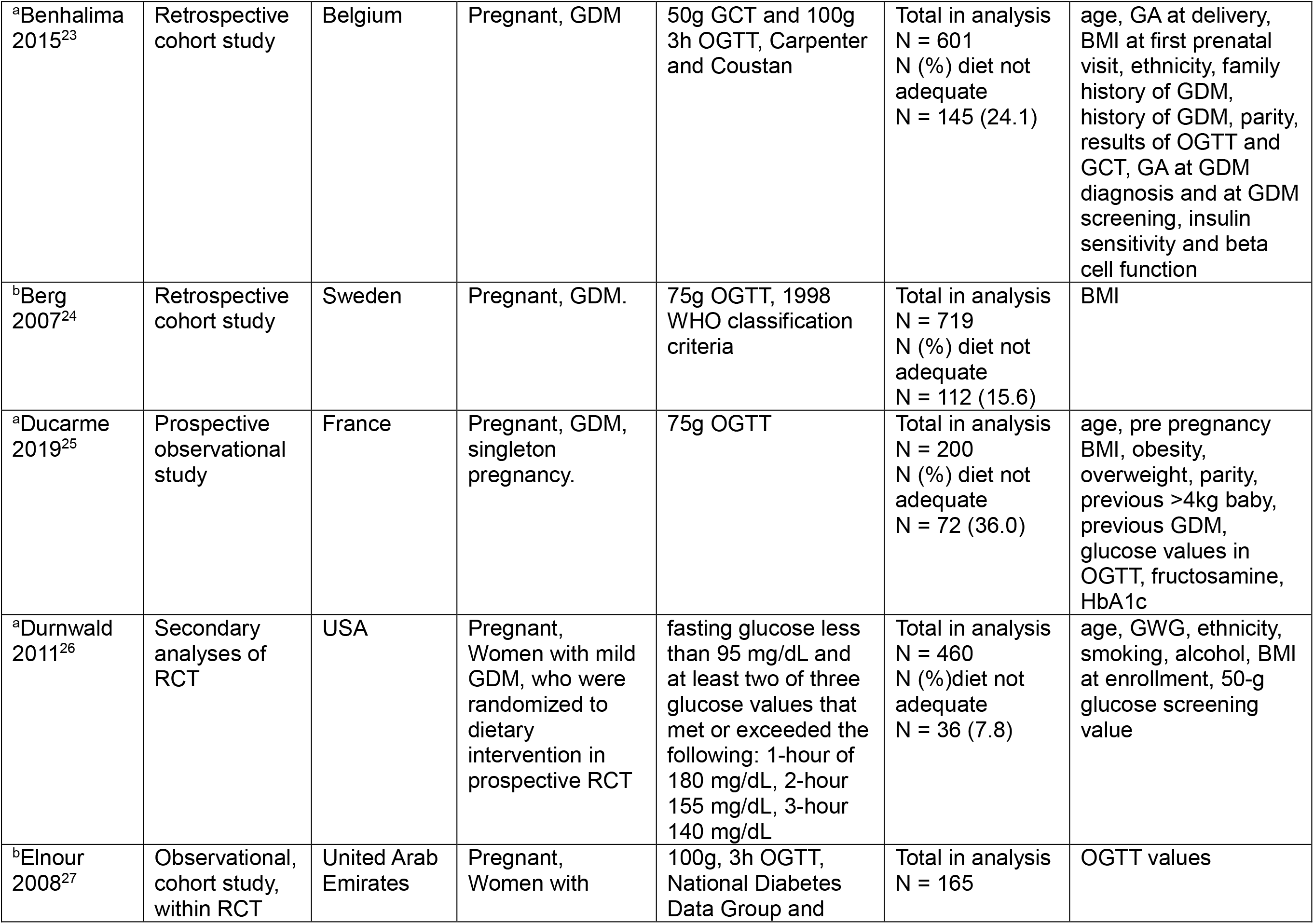
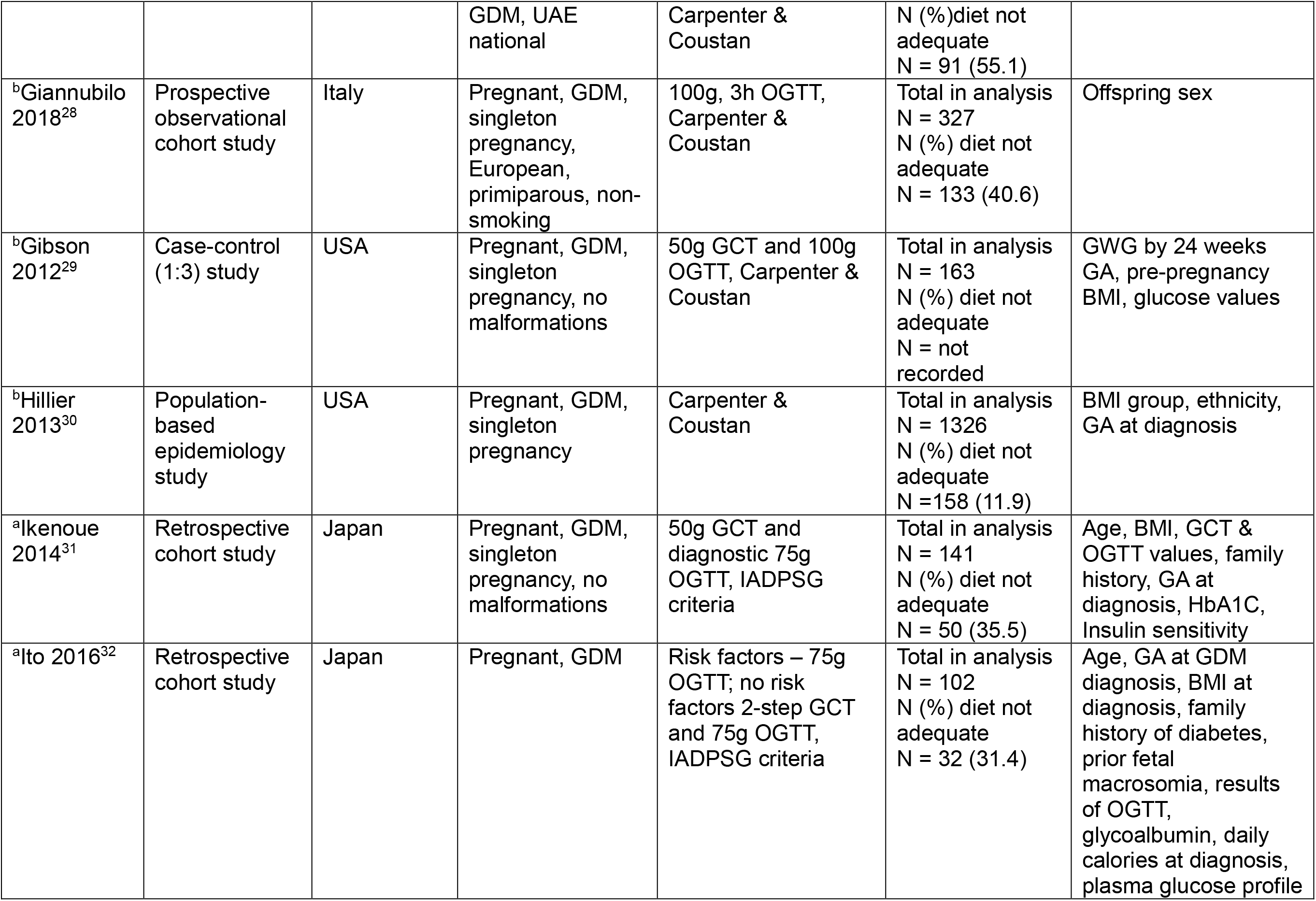
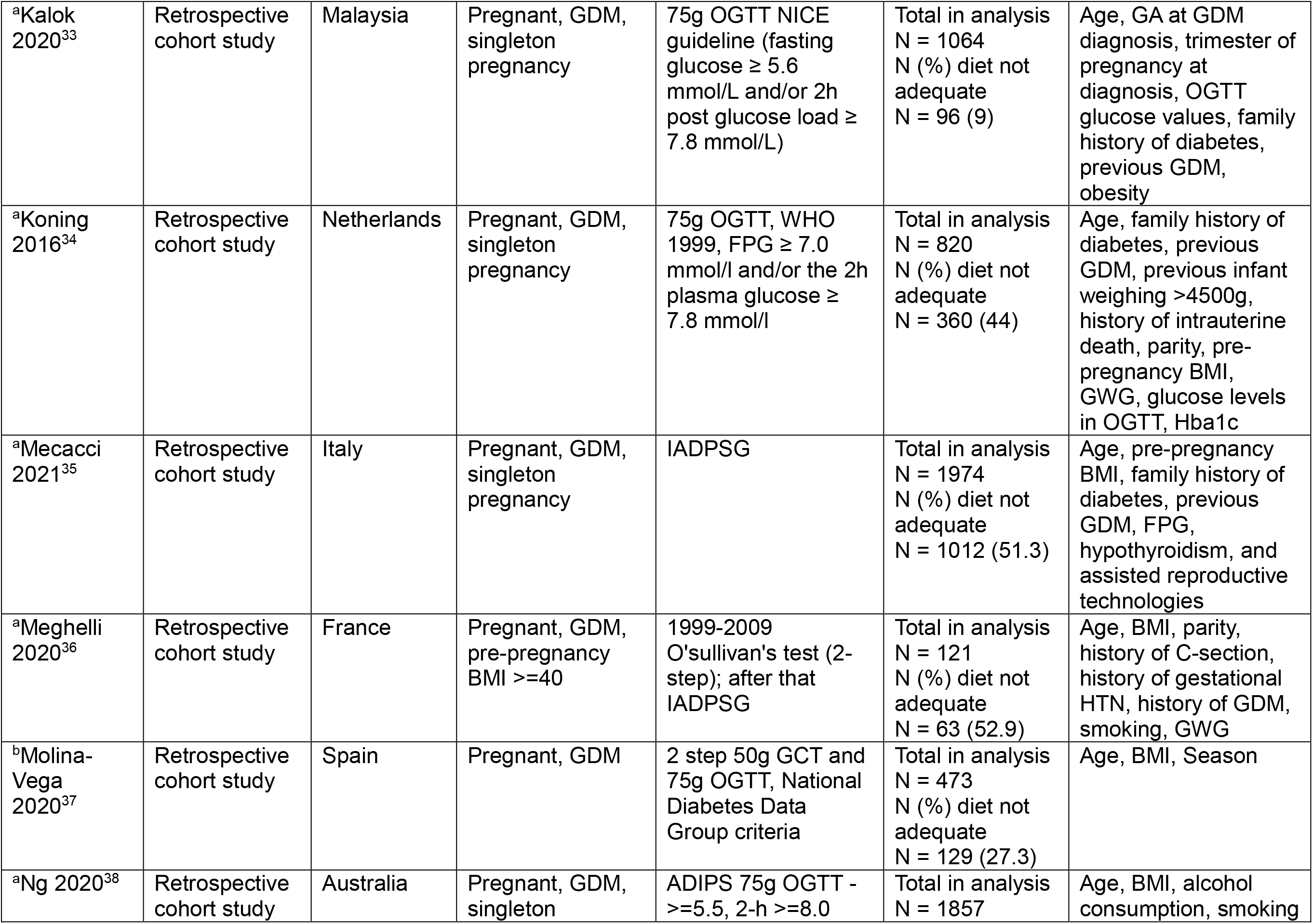
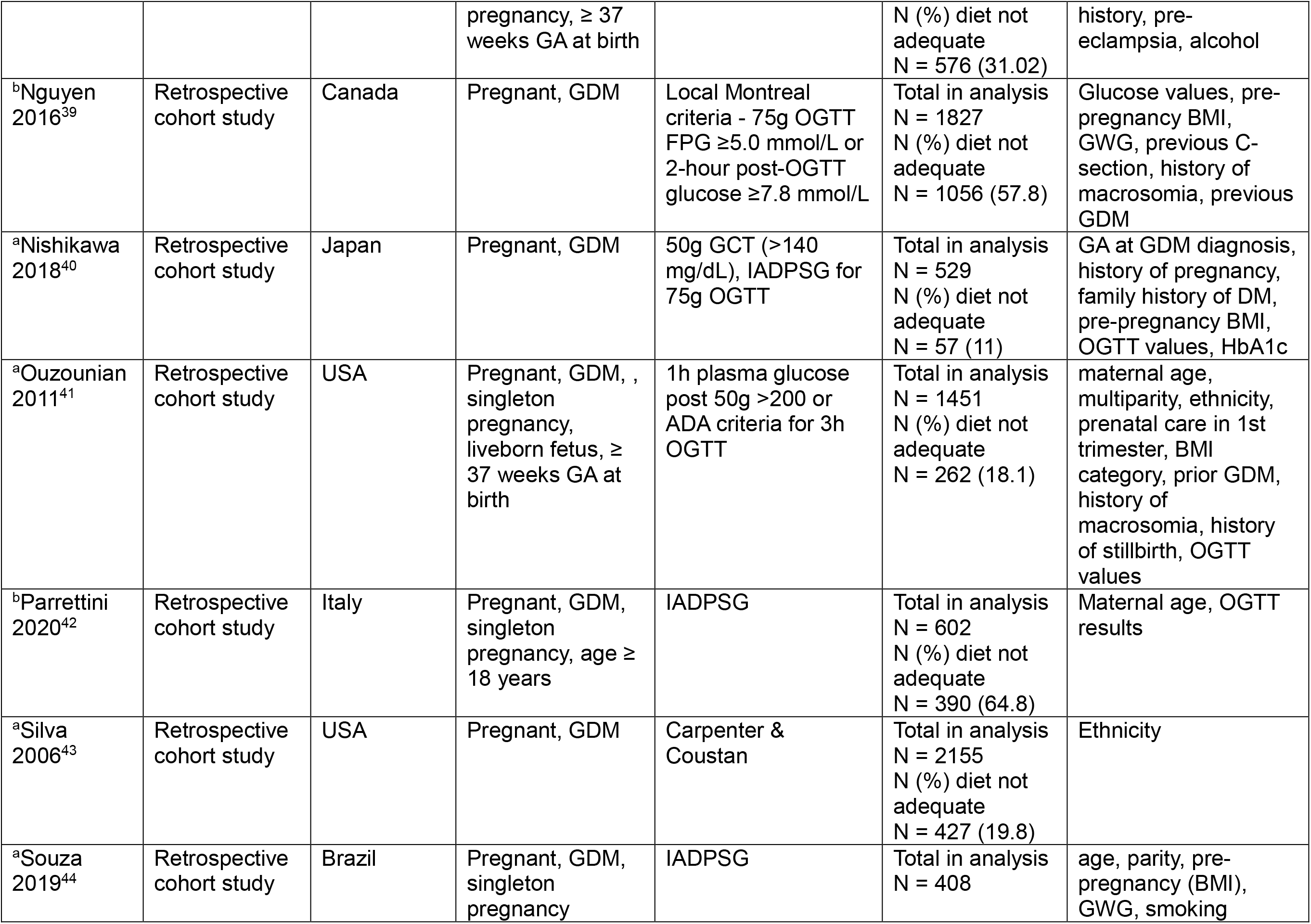
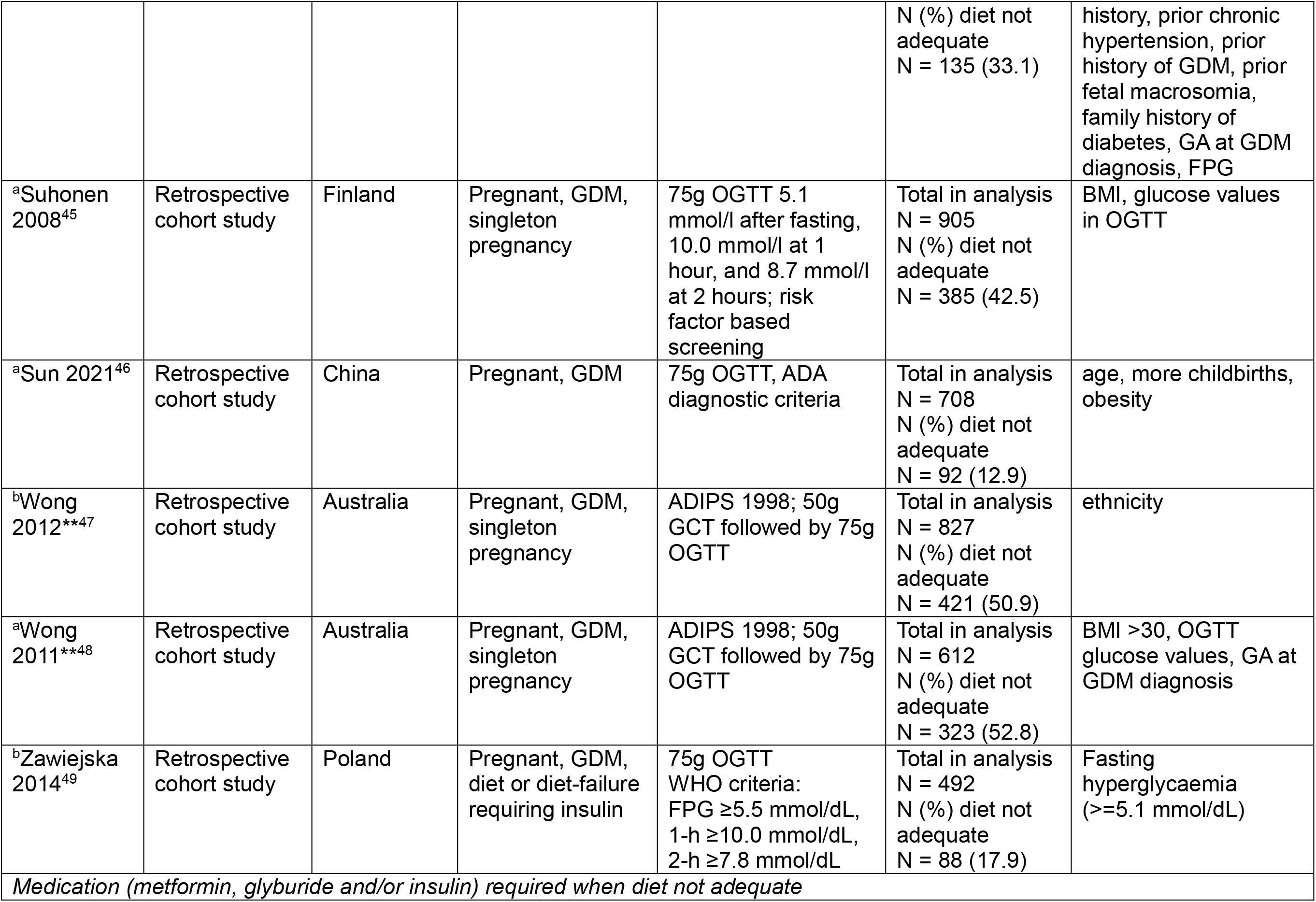
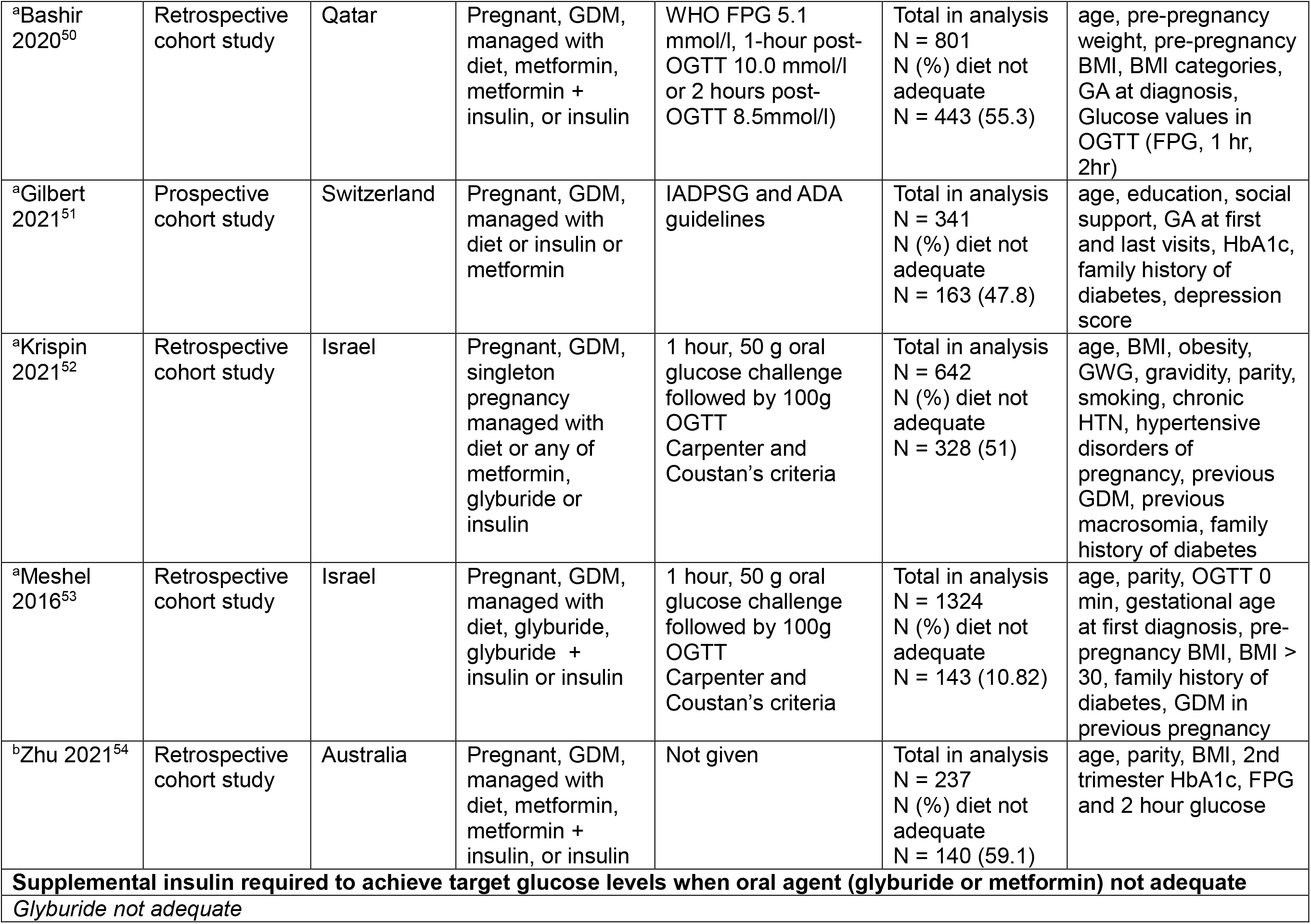
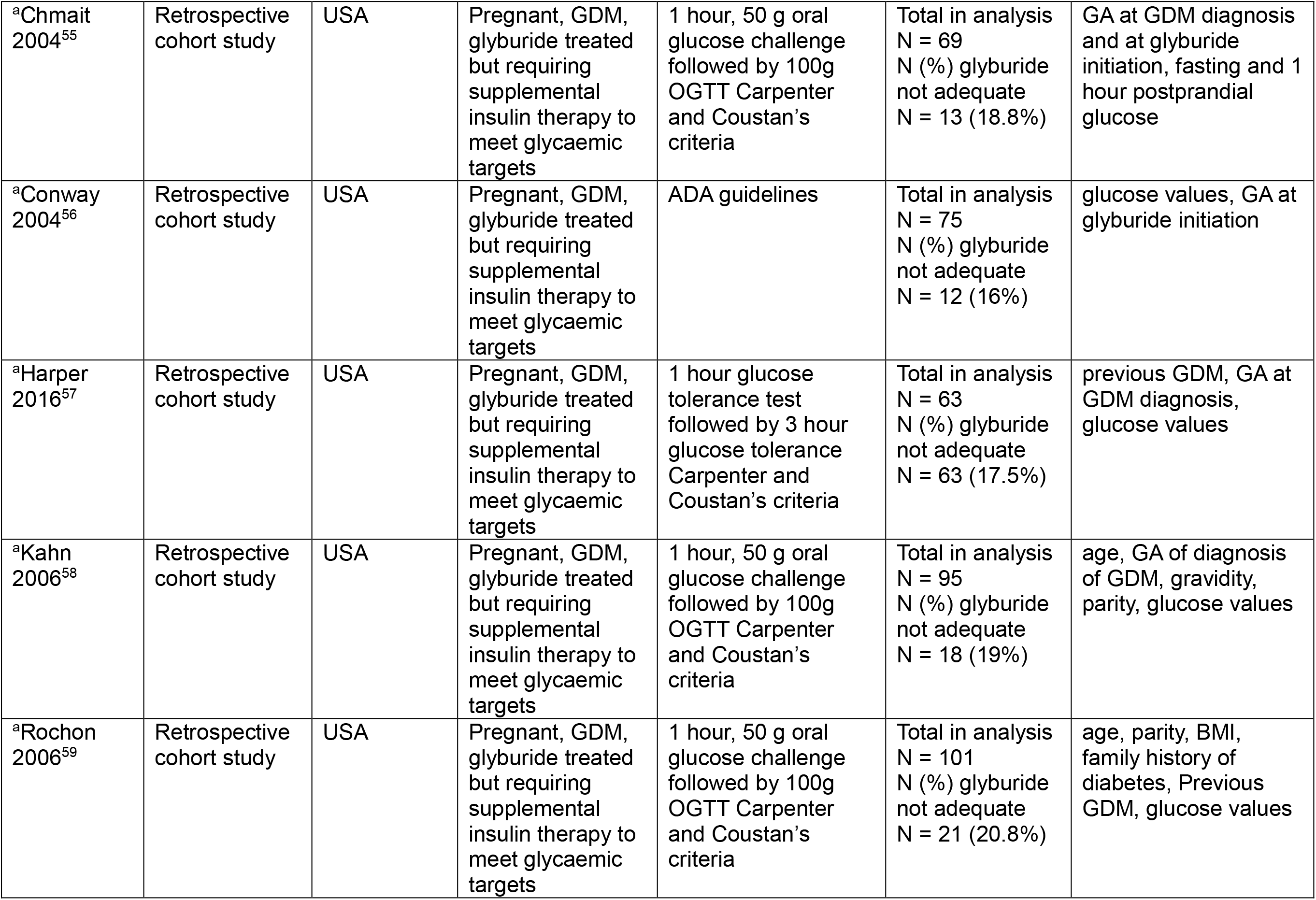
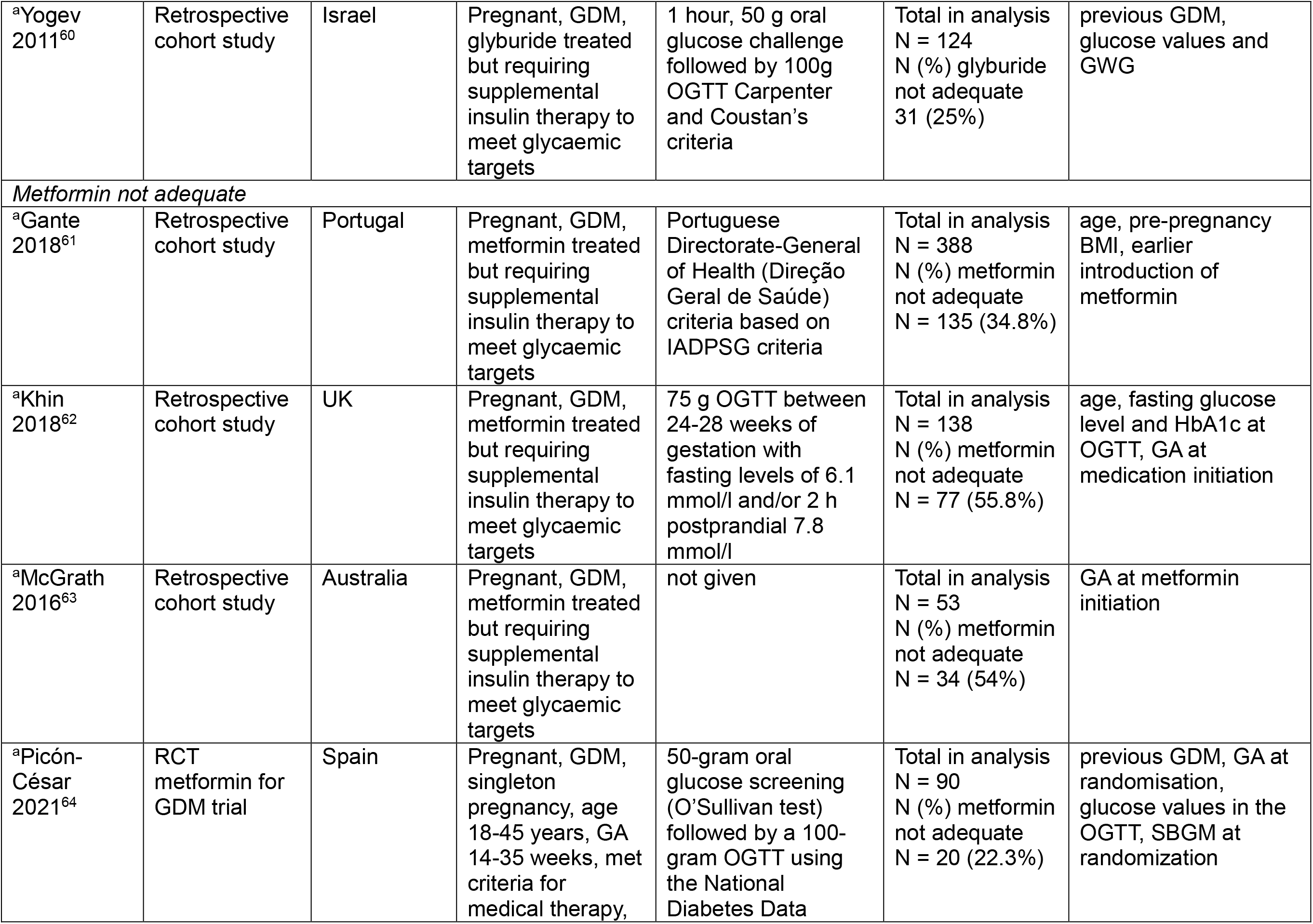
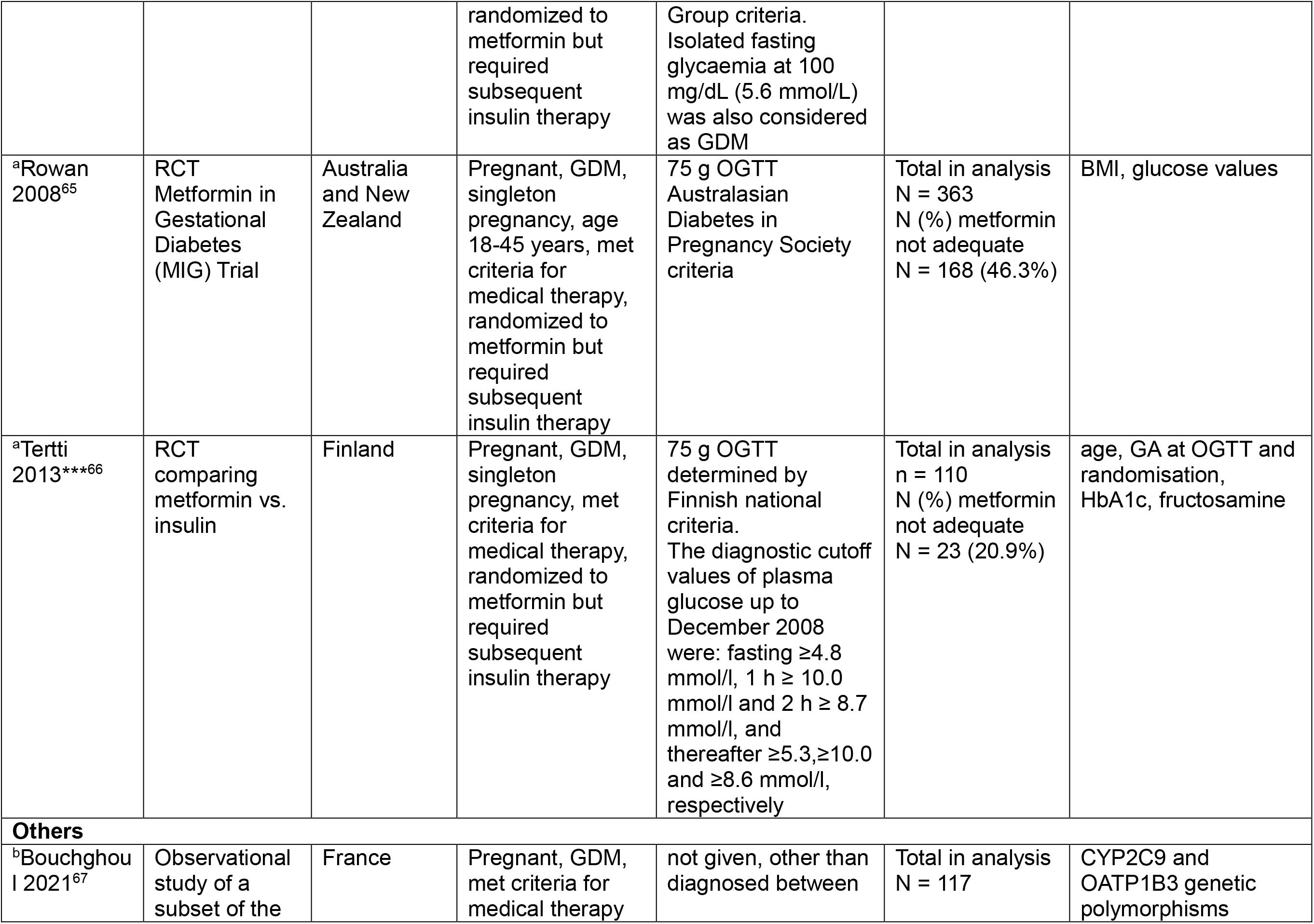
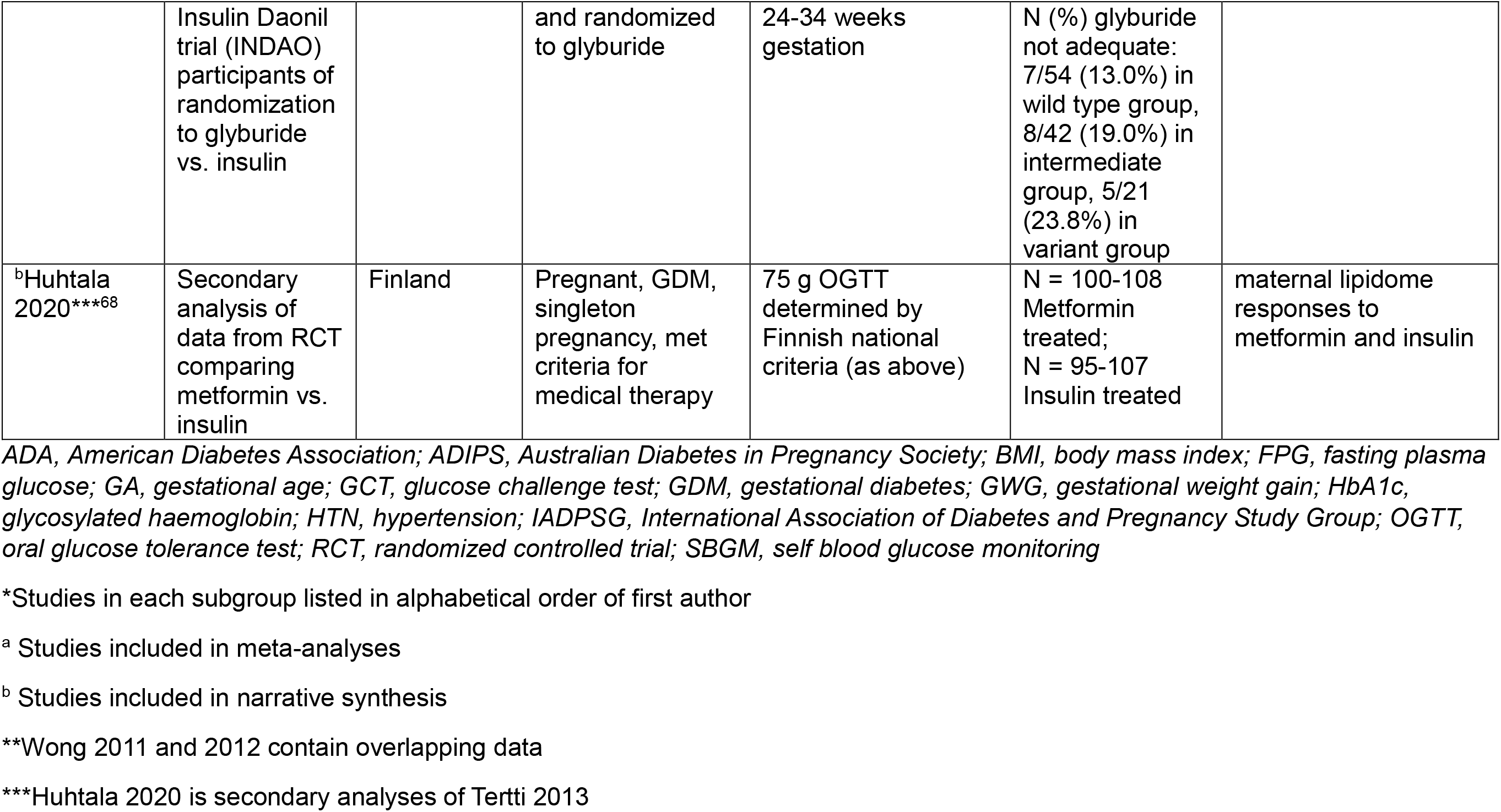
Summary of included studies in the two systematic reviews. Precision predictors of need for pharmacological interventions to achieve target glucose levels

There were 34 studies (n=23,831 participants) where standard care with diet and lifestyle advice was not adequate to achieve target glucose levels. Of these, 29 studies (n=20,486) reported progression to insulin^21-49^ and 5 (n=3,345) reported progression to any medication (metformin, glyburide and/or insulin)^50-54^.

There were 12 studies (n=1,669 participants) where treatment with oral agents was not adequate to achieve target glucose levels, and escalation to insulin was required. Initial treatment was with glyburide in 6 of these studies (n=527)^55-60^ and metformin in the other 6 studies (n=1142)^61-66^.

A further 2 eligible studies reported maternal genetic predictors of need for supplementary insulin after glyburide (n=117 participants) ^67^ and maternal lipidome responses to metformin and insulin (n=217 participants)^68^.

The majority of included studies were observational in design. Most studies reported outcomes of singleton pregnancies. The studies were from a range of geographical locations: Europe (Belgium, Finland, France, Italy, Netherlands, Poland, Portugal, Spain, Sweden), Switzerland, Middle East (Israel, Qatar, United Arab Emirates), Australasia (Australia, New Zealand), North America/Latin America (Canada, USA and Brazil) and Asia (China, Malaysia, Japan). There were a range of approaches to GDM screening, choice of diagnostic test and diagnostic glucose thresholds.

### Quality assessment

Study quality assessment is presented as an overall risk of bias for the studies included in the meta-analyses in Figure 2.1 and as a heat map for quality assessment for each included study in Figure 2.2. Most of the studies were rated as low risk of bias, as they adequately described how a diagnosis of GDM was assigned, defining inclusion and exclusion criteria, and reported the protocol for initiation of pharmacological therapy. Not all studies reported whether women received diet and lifestyle advice as standard care. Few studies reported whether the precision marker was measured in a valid and reliable way. Using the GRADE approach, the majority of precision markers were classified as having a low certainty of evidence with some classified as very low certainty (Tables 2 and 3). No publication bias (as ascertained by funnel plot analyses) was detected.

**Figure 2.1.**
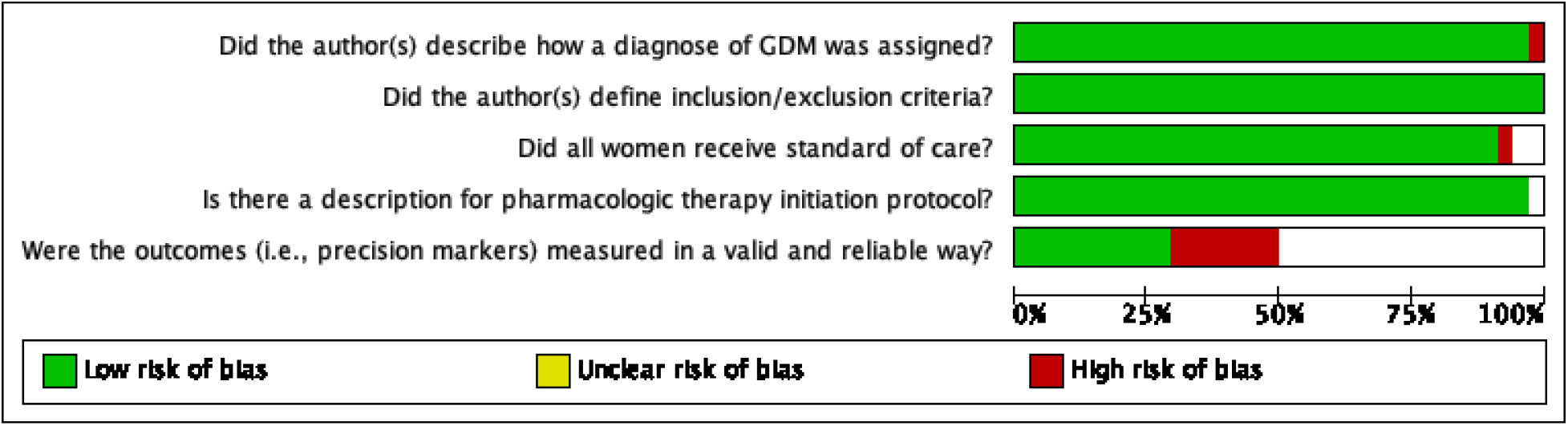
Risk of bias graph: review authors’ judgements about each risk of bias item presented as percentages across all studies included in the meta-analyses.

**Figure 2.2.**
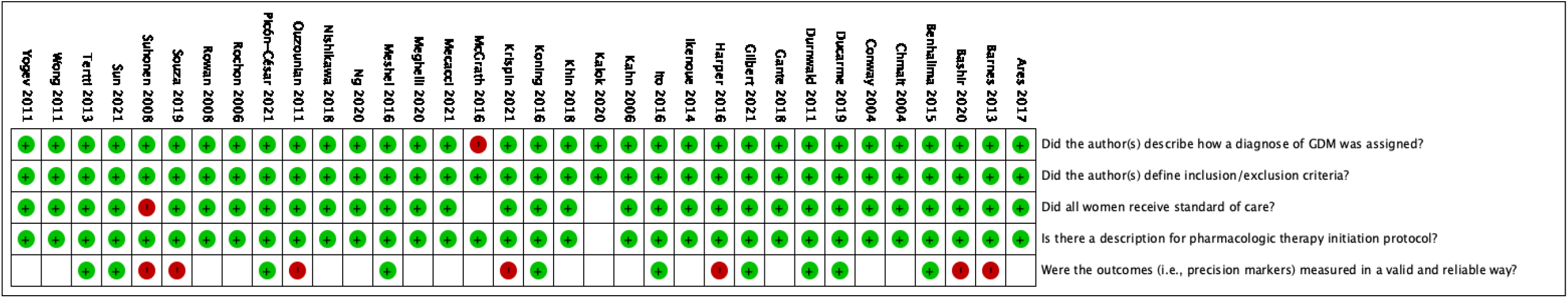
Risk of bias summary: review authors’ judgements about each risk of bias item for each study included in the meta-analyses.

### Synthesis of results

#### Precision diet and lifestyle interventions in GDM

Two studies examining different behavioural interventions were included in the first systematic review, so we present a narrative synthesis of the findings. Neither study examined whether a precision lifestyle intervention enabled achievement of glucose targets during pregnancy.

In one study^19^, the intervention was distribution of a tailored letter detailing gestational weight gain (GWG) recommendations (as defined by the Institute of Medicine). Receipt of this tailored letter increased the likelihood of meeting the end-of-pregnancy weight goal among women with normal pre-pregnancy BMI, but not among women with overweight or obese pre-pregnancy BMI. This study identified normal pre-pregnancy BMI as a precision marker for intervention success.

The second study^20^ used a Web/Smart phone lifestyle coaching program. Pre-intervention excessive GWG was evaluated as a potential precision marker. There was no difference between study arms with respect to either excess GWG or absolute GWG by the end of pregnancy indicating that early GWG is not a useful precision marker with respect to this intervention.

#### Precision predictors of need for pharmacological interventions to achieve glucose targets in GDM

Of the 34 studies of predictors of need for medical therapy in addition to standard care with diet and lifestyle advice to achieve glucose targets, 23 studies (n=19,112 participants) were included in the meta-analysis^21-23,25,26,31-36,38,40,41,43-46,48,50-53^ and 11 studies (n=7158 participants) in the narrative synthesis^24,27-30,37,39,42,47,49,54^.

Table 2 and Supplementary Figures 1.1-1.13 show that precision markers for GDM to be adequately managed with lifestyle measures without need for additional pharmacological therapy were lower maternal age, nulliparity, lower BMI, no previous history of GDM, lower HbA1c, fasting, 1 hour, 2 and 3 hour glucose, no family history of diabetes, later gestation of diagnosis of GDM and no macrosomia in previous pregnancies. There was a similar pattern for not smoking but this did not reach statistical significance.

**Table 2.**
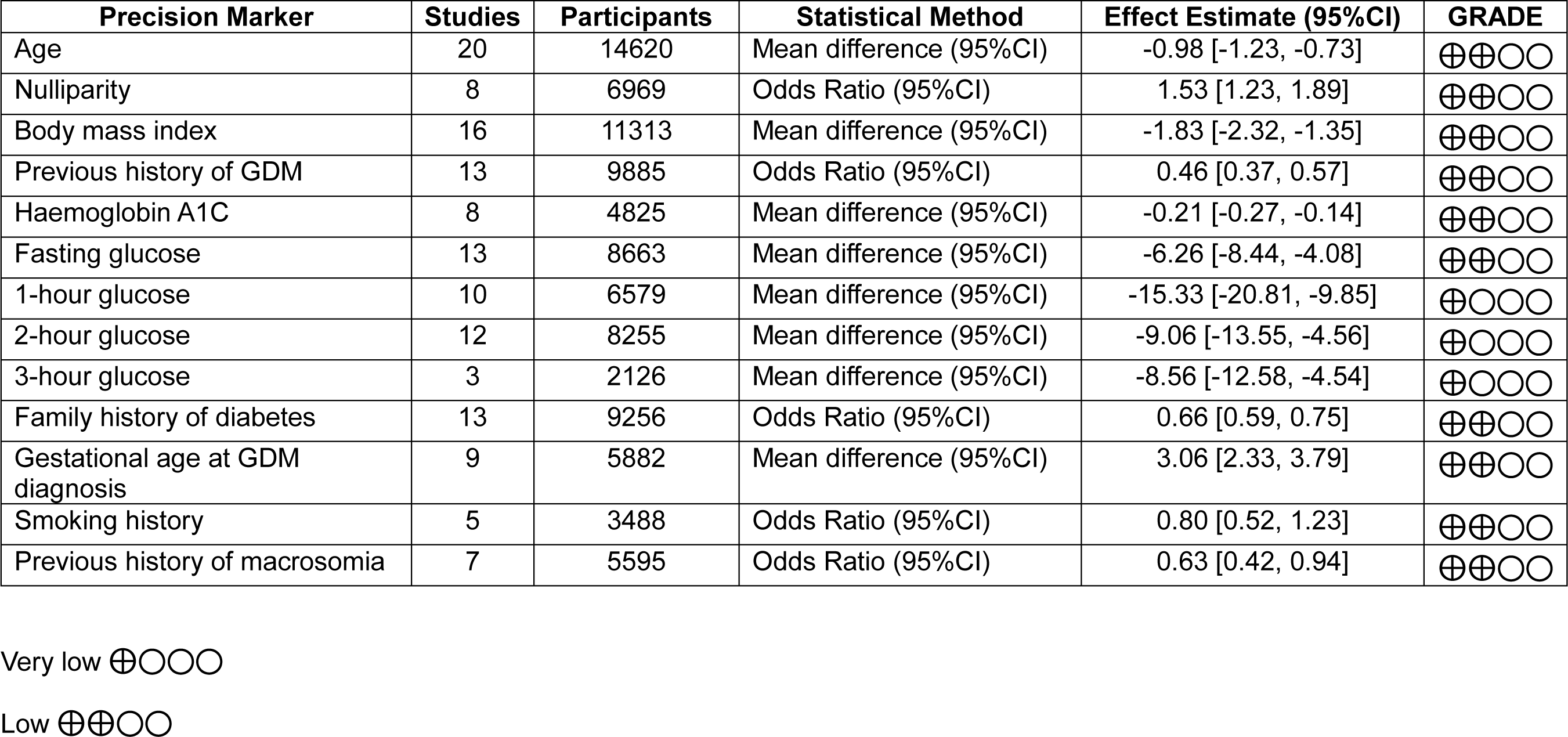
Lifestyle adequate to achieve target glucose levels vs not adequate

| Precision Marker | Studies | Participants | Statistical Method | Effect Estimate (95%CI) | GRADE |
| --- | --- | --- | --- | --- | --- |
| Age | 20 | 14620 | Mean difference (95%CI) | -0.98 [-1.23, -0.73] | ⊕⊕○○ |
| Nulliparity | 8 | 6969 | Odds Ratio (95%CI) | 1.53 [1.23, 1.89] | ⊕⊕○○ |
| Body mass index | 16 | 11313 | Mean difference (95%CI) | -1.83 [-2.32, -1.35] | ⊕⊕○○ |
| Previous history of GDM | 13 | 9885 | Odds Ratio (95%CI) | 0.46 [0.37, 0.57] | ⊕⊕○○ |
| Haemoglobin A1C | 8 | 4825 | Mean difference (95%CI) | -0.21 [-0.27, -0.14] | ⊕⊕○○ |
| Fasting glucose | 13 | 8663 | Mean difference (95%CI) | -6.26 [-8.44, -4.08] | ⊕⊕○○ |
| 1-hour glucose | 10 | 6579 | Mean difference (95%CI) | -15.33 [-20.81, -9.85] | ⊕○○○ |
| 2-hour glucose | 12 | 8255 | Mean difference (95%CI) | -9.06 [-13.55, -4.56] | ⊕○○○ |
| 3-hour glucose | 3 | 2126 | Mean difference (95%CI) | -8.56 [-12.58, -4.54] | ⊕○○○ |
| Family history of diabetes | 13 | 9256 | Odds Ratio (95%CI) | 0.66 [0.59, 0.75] | ⊕⊕○○ |
| Gestational age at GDM diagnosis | 9 | 5882 | Mean difference (95%CI) | 3.06 [2.33, 3.79] | ⊕⊕○○ |
| Smoking history | 5 | 3488 | Odds Ratio (95%CI) | 0.80 [0.52, 1.23] | ⊕⊕○○ |
| Previous history of macrosomia | 7 | 5595 | Odds Ratio (95%CI) | 0.63 [0.42, 0.94] | ⊕⊕○○ |

Very low ⊕○○○
Low ⊕⊕○○

Twelve studies (n=1669 participants) of predictors of need for supplemental insulin to achieve normoglycaemia following treatment with oral agents were included in the meta-analysis^55-66^.

Table 3 and Supplementary Figures 2.1-2.12 show that precision markers for achieving normoglycaemia with oral agents only were nulliparity, lower BMI, no previous history of GDM, lower HbA1c, fasting, 1 hour, and 2 hour glucose, later gestation of diagnosis of GDM and later gestation at initiation of the oral agent. In sensitivity analyses, there were no differences in the precision markers predicting response to metformin versus glyburide.

**Table 3.** Oral pharmacological agent adequate to achieve target glucose levels vs not adequate

| Precision Marker | Studies | Participants | Statistical Method | Effect Estimate (95%CI) | GRADE |
| --- | --- | --- | --- | --- | --- |
| Age | 11 | 1473 | Mean difference (95%CI) | -1.04 [-2.10, 0.03] | ⊕⊕○○ |
| Nulliparity | 8 | 1215 | Odds Ratio (95%CI) | 1.55 [1.17, 2.04] | ⊕⊕○○ |
| Body mass index | 10 | 1692 | Mean difference (95%CI) | -1.21 [-2.21, -0.21] | ⊕⊕○○ |
| Previous history of GDM | 8 | 1412 | Odds Ratio (95%CI) | 0.43 [0.30, 0.63] | ⊕⊕○○ |
| Haemoglobin A1C | 6 | 1152 | Mean difference (95%CI) | -0.21 [-0.29, -0.13] | ⊕⊕○○ |
| Fasting glucose | 12 | 1836 | Mean difference (95%CI) | -8.02 [-11.87, -4.16] | ⊕○○○ |
| 1-hour glucose | 8 | 1177 | Mean difference (95%CI) | -10.64 [-18.25, -3.02] | ⊕○○○ |
| 2-hour glucose | 10 | 1378 | Mean difference (95%CI) | -7.31 [-11.38, -3.25] | ⊕○○○ |
| 3-hour glucose | 6 | 679 | Mean difference (95%CI) | 0.00 [-11.79, 11.79] | ⊕○○○ |
| Family history of diabetes | 6 | 1040 | Odds Ratio (95%CI) | 0.79 [0.50, 1.25] | ⊕⊕○○ |
| Gestational age at GDM diagnosis | 11 | 1473 | Mean difference (95%CI) | 2.64 [1.42, 3.86] | ⊕⊕○○ |
| Gestation at oral pharmacological agent initiation | 7 | 967 | Mean difference (95%CI) | 3.79 [2.08, 5.51] | ⊕⊕○○ |

Very low ⊕○○○
Low ⊕⊕○○

Similar findings were observed in the 11 studies (n=7158 participants) that were not included in the meta-analysis^24,27-30,37,39,42,47,49,54^ (Supplementary Table 1). Additional precision markers including fetal sex^28^, ethnicity^30,47^, and season of birth^37^ were evaluated but there was insufficient data to draw conclusions.

There was a paucity of data in examining other precision markers with only weak evidence that the maternal lipidome^68^ or genetics^67^ hold potential as precision markers of need for pharmacological treatment (Supplementary Table 1).

## DISCUSSION

As the factors contributing to development of GDM are heterogeneous^5-8^, it is plausible that the most effective treatment strategies may also be variable. A precision medicine approach resulting in more rapid normalization of hyperglycaemia could have substantial benefits for both mother and fetus. By synthesizing the evidence from two systematic reviews, we sought to identify key precision markers that may predict effective lifestyle and pharmacological interventions. There were a paucity of studies examining precision lifestyle-based interventions for GDM highlighting the pressing need for further research in this area. However, we found a number of precision markers to enable earlier identification of those requiring escalation of pharmacological therapy. These included characteristics such as BMI, that are easily and routinely measured in clinical practice, and thus have potential to be integrated into prediction models with the aim of achieving rapid glycaemic control. With the relatively short timeframe available to treat GDM, commencing effective therapy earlier, and thus reducing excess fetal growth, is an important target to improve outcomes. Basing treatment decisions closely on precision markers could also avoid over-medicalisation of women who are likely to achieve glucose targets with dietary counselling alone.

In our first systematic review we identified only two studies addressing precision markers in lifestyle-based interventions for GDM, over and above standard care^19,20^. In both studies, precision markers were examined as secondary analyses of the trials and only two precision markers (BMI and GWG) were assessed; it is thus not possible to conclusively identify any precision marker in lifestyle-based interventions for GDM. This gap in the literature highlights the need for more research, as also echoed by patients and healthcare professionals participating in the 2020 James Lind Alliance (JLA) Priority Setting Partnership (PSP)^69^.

Our second systematic review extends the observations of a previous systematic review reporting maternal characteristics associated with the need for insulin treatment in GDM^11^. We identified a number of additional precision markers of successful GDM treatment with lifestyle measures alone, without need for additional pharmacological therapy. The same set of predictors identified women requiring additional insulin after treatment with glyburide as with metformin, despite their different mechanisms of action. However the numbers of women included in most studies were relatively low and most studies with data in relation to glyburide failure were over 10 years old^55,56,58-60^. We acknowledge that there are also differences in diagnostic criteria, clinical practices, and preferences for choice of which drug to start as first pharmacological agent in various global regions which may limit the generalizability of our findings.

Notably, many of the identified precision markers are routinely measured in clinical practice and so could be incorporated into prediction models of need for pharmacological treatment^70,71^. By identifying those who require escalation of pharmacological therapy earlier, better allocation of resources can be achieved. Additionally, some of the precision markers identified, such as BMI, are potentially modifiable. This raises the question of how women can be helped to better prepare for pregnancy^72^. Implementing interventions prior to pregnancy could help understand if these precision markers are on the causal pathway, thus providing an opportunity for prevention and improving health outcomes.

Importantly, there was a lack of data on other potential precision treatment biomarkers, with only two eligible low quality studies reporting maternal genetic and metabolomic findings^67,68^. In the non-pregnancy literature, efficacy of dietary interventions has been reported to differ for patients with distinct metabolic profiles, for example high fasting glucose vs high fasting insulin, or insulin resistance vs low insulin secretion^73-75^. More recent evidence from appropriately designed, prospective dietary intervention studies has confirmed that dietary interventions tailored towards specific metabolic profiles have more beneficial effects than interventions not specifically designed towards a patient’s metabolic profile^76-79^. Ongoing studies such as the Westlake Precision Birth Cohort (WeBirth) in China (NCT04060056) and the USA Hoosier Moms Cohort (NCT03696368) are collecting additional biomarkers which will enhance knowledge in this field. However implementing such measures in clinical practice, if they prove informative, could be complex and expensive and thus not suitable for use in all global contexts.

Our study has several limitations: Our reviews primarily relied on secondary analyses from observational studies that were not specifically designed to address the question of precision medicine in GDM treatment and were not powered for many of the comparisons made. Prior to introduction in clinical practice, any marker would have to be rigorously and prospectively tested with respect to sensitivity and specificity to predict treatment needs. The majority of data were extracted from clinical records leading to a lack of detail, such as the precise timing of BMI measurements, and limited information about whether BMI was self-reported or clinician measured. There was marked variation in approaches to GDM screening methods, choice of glucose challenge test and diagnostic thresholds. Whilst we included studies from a range of geographical settings, the majority of studies were from high income settings, and therefore our findings may not be applicable to low- and middle-income countries. Pregnancy outcomes of precision medicine strategies for GDM also remain unknown, underscoring the need for tailored interventions that account for patient perspective and diverse patient populations.

Despite these limitations, our study has several strengths. We used robust methods to identify a broad range of precision markers, many of which are routinely measured and can be easily translated into prediction models. We excluded studies where the choice of drug was decided by the clinician based on participant characteristics to avoid bias. Our study also highlights the need for further research in this area, particularly in exploring whether there are more sensitive markers that could be identified through “omics” approaches.

In conclusion, our findings suggest that precision medicine for GDM treatment holds promise as a tool to stream-line individuals towards the most effective and potentially cost-effective care. Whether this will impact on short-term pregnancy outcomes and longer term health outcomes for both mother and baby is not known. More research is urgently needed to identify precision lifestyle interventions and to explore whether more sensitive markers could be identified. Prospective studies, appropriately powered and designed to allow assessment of discriminative abilities (sensitivity, specificity), and (external) validation studies are urgently needed to understand the utility and generalizability of our findings to under-represented populations. Consideration of how identified markers can be implemented feasibly and cost effectively in clinical practice is also required. Such efforts will be critical for realising the full potential of precision medicine and empowering patients and their health care providers to optimise short and long-term health outcomes for both mother and child.

## Author contributions

All authors contributed to the design of the research questions, study selection, extraction of data, data analyses, quality assessment and data interpretation. RMR wrote the first draft of the manuscript. All authors edited the manuscript and all approved the final version.

## Conflicts of Interest

None of the authors have any conflicts of interest to declare.

## Funding

The ADA/EASD Precision Diabetes Medicine Initiative, within which this work was conducted, has received the following support: The Covidence license was funded by Lund University (Sweden) for which technical support was provided by Maria Björklund and Krister Aronsson (Faculty of Medicine Library, Lund University, Sweden). Administrative support was provided by Lund University (Malmö, Sweden), University of Chicago (IL, USA), and the American Diabetes Association (Washington D.C., USA). The Novo Nordisk Foundation (Hellerup, Denmark) provided grant support for in-person writing group meetings (PI: L Phillipson, University of Chicago, IL). JMM acknowledges the support of the Henry Friesen Professorship in Endocrinology, University of Manitoba, Canada. NMM and RMR acknowledge the support of the British Heart Foundation (RE/18/5/34216). SEO is supported by the Medical Research Council (MC_UU_00014/4) and British Heart Foundation (RG/17/12/33167).

## Data Availability

All data produced in the present study are available upon reasonable request to the authors

## Notes

### Competing Interest Statement

The authors have declared no competing interest.

### Clinical Protocols

https://www.crd.york.ac.uk/prospero/display_record.php?RecordID=299288

https://www.crd.york.ac.uk/prospero/display_record.php?RecordID=299402

