## Supplementary material for "Precision Gestational Diabetes Treatment: Systematic review and Meta-analyses"

**Supplementary Text S1 Search Strategy**

**Supplementary Figures 1.1 to 1.13 Forest Plots (A) and Funnel Plots (B) for GDM to be adequately managed with lifestyle measures without need for additional pharmacological therapy**

**Supplementary Figures 2.1 to 2.12 Forest Plots (A) and Funnel Plots (B) for Oral Pharmacological Agent adequate in controlling glucose vs not adequate**

**Supplementary Table 1 Narrative summary of studies not included in the meta-analysis**

**Supplementary Text S1 Search Strategy**

**PubMed Search**

**Systematic review 1: Lifestyle interventions**

#1 Diabetes, Gestational"[Mesh]

#2 gestational diabetes"[Title/Abstract] OR GDM[Title/Abstract] OR pregnancy induced diabetes[Title/Abstract] OR pregnancy-induced diabetes

#3 #1 OR #2

a.#4 ((((("Body-Weight Trajectory"[Mesh]) OR "Body Mass Index"[Mesh]) OR "Body Weight"[Mesh]) OR "Body Composition"[Mesh]) OR "Waist Circumference"[Mesh]) OR weight gain[MeSH]

#5 bodyweight*[Title/Abstract] OR "body weight*"[Title/Abstract] OR body-weight*[Title/Abstract] OR bmi[Title/Abstract] OR "body mass index"[Title/Abstract] OR "body composition"[Title/Abstract] OR bodycomposition[Title/Abstract] OR body-composition[Title/Abstract] OR "body fat"[Title/Abstract] OR bodyfat body-fat[Title/Abstract] OR "waist circumfer*"[Title/Abstract] OR waistcircumfer*[Title/Abstract] OR waist-circumfer*[Title/Abstract] OR "weight gain"[Title/Abstract] OR weight-gain[Title/Abstract] OR weightgain[Title/Abstract]

#6 #4 OR #5

b. #7 ((("Diet"[Mesh]) OR "Dietary Supplements"[Mesh]) OR "Diet, Carbohydrate-Restricted"[Mesh]) OR "Diet, Fat-Restricted"[Mesh]) OR "Caloric Restriction"[Mesh] OR nutrition[MeSH]

#8 diet*[Title/Abstract] OR "caloric restrict*"[Title/Abstract] OR calory-restrict*[Title/Abstract] OR eating[Title/Abstract] OR macronutri*[Title/Abstract] OR nutrition*[Title/Abstract] OR protein*[Title/Abstract] OR meal[Title/Abstract] OR beverage*[Title/Abstract] OR meat*[Title/Abstract] OR behavior*[Title/Abstract] OR behaviour*[Title/Abstract] OR habit*[Title/Abstract] OR sleep*[Title/Abstract] OR food*[Title/Abstract]

#9 #7 OR #8

c. #10 exercise[MeSH] OR physical fitness [MeSH] OR lifestyle [MeSH] OR healthy lifestyle [MeSH] OR sedentary behavior [MeSH]

#11 exercis*[Title/Abstract] OR "physical activit*"[Title/Abstract] OR fitness[Title/Abstract] OR sedentary[Title/Abstract] OR walk*[Title/Abstract] OR stretch*[Title/Abstract] OR lifestyle*[Title/Abstract] OR "life style*"[Title/Abstract] OR life-style*[Title/Abstract] OR wellness[Title/Abstract] OR "strength train*"[Title/Abstract] OR strength-train*[Title/Abstract]

#12 #10 OR #11

#13 #6 OR #9 OR #12

#14 #3 AND #13

#15 ("controlled trial*" OR randomi* OR "observational stud*" OR RCT OR "retrospective stud*" OR Metformin in Gestational Diabetes trial ) OR ("controlled trial*"[Publication Type] OR randomi*[Publication Type] OR "observational stud*"[Publication Type] OR RCT[Publication Type] OR "retrospective stud*"[Publication Type])

#16 #14 AND #15

#17 #16 filters: humans, English

**Systematic review 2: Pharmacological interventions**

#1 “Diabetes, Gestational"[Mesh]

#2 "gestational diabetes"[Title/Abstract] OR GDM[Title/Abstract] OR pregnancy induced diabetes[Title/Abstract] OR pregnancy-induced diabetes

#3 #1 OR #2

#4 "Insulin"[Mesh]

#5 ((("Metformin"[Mesh]) OR "Sulfonylurea Compounds"[Mesh]) OR "Glyburide"[Mesh]) OR "Secretagogues"[Mesh]

#6 insulin*[Title/Abstract] OR novolin[Title/Abstract] OR iletin[Title/Abstract] OR sulfonylurea*[Title/Abstract] OR Acetohexamide[Title/Abstract] OR Carbutamide[Title/Abstract] OR Chlorpropamide[Title/Abstract] OR Gliclazide[Title/Abstract] OR Glyburide[Title/Abstract] OR Tolazamide[Title/Abstract] OR Tolbutamide[Title/Abstract] OR sulphonylurea*[Title/Abstract] OR glibenclamide[Title/Abstract] OR secretagogues[Title/Abstract] OR “pharmacological therapy”[Title/Abstract]

#7 #4 OR #5 OR #6

#8 #3 AND #7

#9 ("controlled trial*" OR randomi* OR "observational stud*" OR RCT OR "retrospective stud*" OR Metformin in Gestational Diabetes trial ) OR ("controlled trial*"[Publication Type] OR randomi*[Publication Type] OR "observational stud*"[Publication Type] OR RCT[Publication Type] OR "retrospective stud*"[Publication Type])

#10 #8 AND #9

#11 #10 Filters: humans, English

**Embase search**

#1 'pregnancy diabetes mellitus'/exp

#2 'gestational diabetes':ab,ti OR gdm:ab,ti OR 'pregnancy-induced diabetes':ab,ti OR 'pregnancy induced diabetes':ab,ti

#3 #1 OR #2

**Systematic review 1: Lifestyle interventions**

#4 'weight trajectory' AND 'body weight'/exp OR 'body mass'/exp OR 'body weight'/exp OR 'body composition'/exp OR 'waist circumference'/exp OR 'body weight gain'/exp

#5 bodyweight*:ab,ti OR 'body weight*':ab,ti OR 'body-weight*or bmi':ab,ti OR 'body mass index':ab,ti OR bodycomposition:ab,ti OR 'body composition':ab,ti OR 'body fator body-fat':ab,ti OR 'waist circumfer*':ab,ti OR 'waistcircumfer*or waist-circumfer*':ab,ti OR 'weight gain':ab,ti OR weightgain:ab,ti

#6 #4 OR #5

#7 'diet'/exp OR 'dietary supplement'/exp OR 'low carbohydrate diet'/exp OR 'low fat diet'/exp OR 'caloric restriction'/exp OR 'nutrition'/exp

#8 diet*:ab,ti OR 'caloric restrict*':ab,ti OR 'calory restrict*':ab,ti OR eating:ab,ti OR macronutri*:ab,ti OR nutrition*:ab,ti OR protein*:ab,ti OR meal:ab,ti OR beverage*:ab,ti OR meat*:ab,ti OR behavior*:ab,ti OR behaviour*:ab,ti OR habit*:ab,ti OR sleep*:ab,ti OR food*:ab,ti

#9 #7 OR #8

#10 'exercise'/exp OR 'fitness'/exp OR 'lifestyle'/exp OR 'healthy lifestyle'/exp OR 'sedentary lifestyle'/exp

#11 exercis*:ab,ti OR 'physical activit*':ab,ti OR fitness:ab,ti OR sedentary:ab,ti OR walk*:ab,ti OR stretch*:ab,ti OR lifestyle*:ab,ti OR 'life style*':ab,ti OR wellness:ab,ti OR 'strength train*':ab,ti

#12 #10 OR #11

#13 #6 OR #9 OR #12

**Study design filter**

#14 ((randomized:ab,ti OR randomised:ab,ti OR randomly:ab,ti OR rct:ab,ti OR 'retrospective stud*':ab,ti OR controlled) AND clinical AND trial:ab,ti OR controlled) AND trial:ab,ti OR 'randomized controlled trial'/de OR 'controlled clinical trial'/de

Combination search RQ1 Lifestyle interventions

GDM + Lifestyle interventions + study design filter

#15 #3 AND #13 AND #14

AND [embase]/lim NOT ([embase]/lim AND [medline]/lim)

Filters Human, English

NOT 'conference abstract':it

**Systematic review 2: Pharmacological interventions**

#16 'insulin'/exp

#17 'metformin'/exp OR 'sulfonylurea derivative'/exp OR 'glibenclamide'/exp OR 'secretagogue'/exp

#18 (insulin* OR novolin OR iletin OR sulfonylurea* OR acetohexamide OR carbutamide OR chlorpropamide OR gliclazide OR glyburide OR tolazamide OR tolbutamide OR sulphonylurea* OR glibenclamide OR secretagogues OR pharmacological) AND therapy

#19 #16 OR #17 OR #18

Combination search RQ2 Pharmacological interventions

#6 GDM + Pharmacological interventions + study design filter

#3 AND #19 AND #13

AND [embase]/lim NOT ([embase]/lim AND [medline]/lim)

Filters Human, English

NOT 'conference abstract':it

**Supplementary Figures 1.1 to 1.13**

**Forest Plots (A) and Funnel Plots (B) for GDM to be adequately managed with lifestyle measures without need for additional pharmacological therapy**

**Supplementary Figure 1.1A** Forest plot for included studies comparing if lifestyle was adequate or not adequate for maternal age

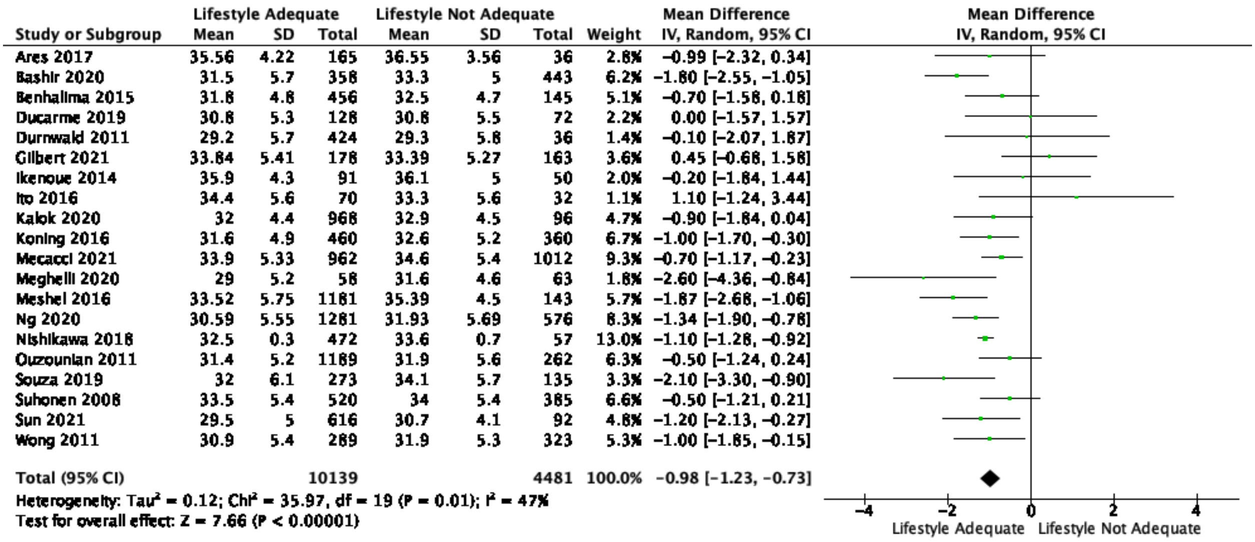

| Study or Subgroup | Lifestyle Adequate Mean | SD | Total | Lifestyle Not Adequate Mean | SD | Total | Weight | Mean Difference IV, Random, 95% CI |
|---|---|---|---|---|---|---|---|---|
| Ares 2017 | 35.56 | 4.22 | 165 | 36.55 | 3.56 | 36 | 2.8% | −0.99 [−2.32, 0.34] |
| Bashir 2020 | 31.5 | 5.7 | 358 | 33.3 | 5 | 443 | 6.2% | −1.80 [−2.55, −1.05] |
| Benhalima 2015 | 31.8 | 4.8 | 456 | 32.5 | 4.7 | 145 | 5.1% | −0.70 [−1.58, 0.18] |
| Ducarme 2019 | 30.8 | 5.3 | 128 | 30.8 | 5.5 | 72 | 2.2% | 0.00 [−1.57, 1.57] |
| Durnwald 2011 | 29.2 | 5.7 | 424 | 29.3 | 5.8 | 36 | 1.4% | −0.10 [−2.07, 1.87] |
| Gilbert 2021 | 33.84 | 5.41 | 178 | 33.39 | 5.27 | 163 | 3.6% | 0.45 [−0.68, 1.58] |
| Ikenoue 2014 | 35.9 | 4.3 | 91 | 36.1 | 5 | 50 | 2.0% | −0.20 [−1.84, 1.44] |
| Ito 2016 | 34.4 | 5.6 | 70 | 33.3 | 5.6 | 32 | 1.1% | 1.10 [−1.24, 3.44] |
| Kalok 2020 | 32 | 4.4 | 968 | 32.9 | 4.5 | 96 | 4.7% | −0.90 [−1.84, 0.04] |
| Koning 2016 | 31.6 | 4.9 | 460 | 32.6 | 5.2 | 360 | 6.7% | −1.00 [−1.70, −0.30] |
| Mecacci 2021 | 33.9 | 5.33 | 962 | 34.6 | 5.4 | 1012 | 9.3% | −0.70 [−1.17, −0.23] |
| Meghelli 2020 | 29 | 5.2 | 58 | 31.6 | 4.6 | 63 | 1.8% | −2.60 [−4.36, −0.84] |
| Meshel 2016 | 33.52 | 5.75 | 1181 | 35.39 | 4.5 | 143 | 5.7% | −1.87 [−2.68, −1.06] |
| Ng 2020 | 30.59 | 5.55 | 1281 | 31.93 | 5.69 | 576 | 8.3% | −1.34 [−1.90, −0.78] |
| Nishikawa 2018 | 32.5 | 0.3 | 472 | 33.6 | 0.7 | 57 | 13.0% | −1.10 [−1.28, −0.92] |
| Ouzounian 2011 | 31.4 | 5.2 | 1189 | 31.9 | 5.6 | 262 | 6.3% | −0.50 [−1.24, 0.24] |
| Souza 2019 | 32 | 6.1 | 273 | 34.1 | 5.7 | 135 | 3.3% | −2.10 [−3.30, −0.90] |
| Suhonen 2008 | 33.5 | 5.4 | 520 | 34 | 5.4 | 385 | 6.6% | −0.50 [−1.21, 0.21] |
| Sun 2021 | 29.5 | 5 | 616 | 30.7 | 4.1 | 92 | 4.8% | −1.20 [−2.13, −0.27] |
| Wong 2011 | 30.9 | 5.4 | 289 | 31.9 | 5.3 | 323 | 5.3% | −1.00 [−1.85, −0.15] |
| Total (95% CI) | | | 10139 | | | 4481 | 100.0% | −0.98 [−1.23, −0.73] |

Heterogeneity: Tau² = 0.12; Chi² = 35.97, df = 19 (P = 0.01); I² = 47%
Test for overall effect: Z = 7.66 (P < 0.00001)



**Supplementary Figure 1.1B** Funnel Plot for Assessment of Publication Bias

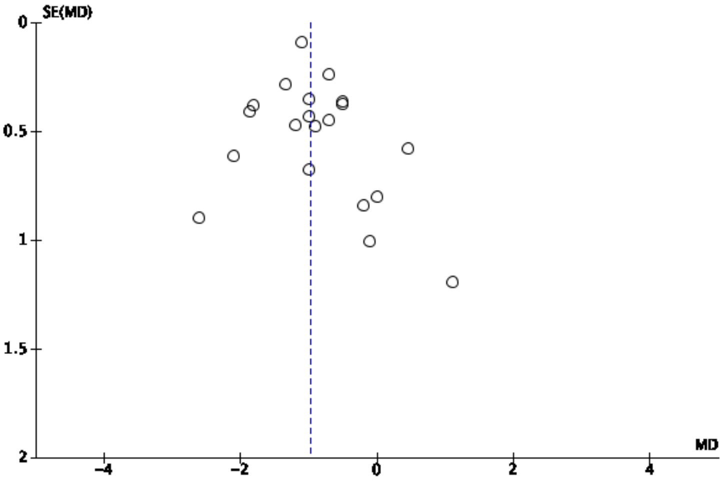

**Supplementary Figure 1.2A** Forest plot for included studies comparing if lifestyle was adequate or not adequate for nulliparity

| Study or Subgroup | Lifestyle Adequate Events | Lifestyle Adequate Total | Lifestyle Not Adequate Events | Lifestyle Not Adequate Total | Weight | Odds Ratio M-H, Random, 95% CI |
|---|---|---|---|---|---|---|
| Benhalima 2015 | 163 | 456 | 34 | 145 | 10.9% | 1.82 [1.18, 2.79] |
| Ducarme 2019 | 48 | 126 | 23 | 72 | 7.5% | 1.31 [0.71, 2.42] |
| Koning 2016 | 223 | 460 | 110 | 360 | 14.3% | 2.14 [1.60, 2.86] |
| Mecacci 2021 | 590 | 962 | 578 | 1012 | 17.0% | 1.19 [0.99, 1.43] |
| Ouzounian 2011 | 497 | 1189 | 77 | 262 | 14.3% | 1.73 [1.29, 2.31] |
| Souza 2019 | 74 | 273 | 24 | 135 | 9.1% | 1.72 [1.03, 2.88] |
| Suhonen 2008 | 187 | 520 | 91 | 385 | 14.1% | 1.81 [1.35, 2.44] |
| Wong 2011 | 85 | 289 | 101 | 323 | 12.9% | 0.92 [0.65, 1.29] |
| Total (95% CI) | | 4275 | | 2694 | 100.0% | 1.53 [1.23, 1.89] |
| Total events | 1867 | | 1038 | | | |

Heterogeneity: $Tau^2 = 0.06$; $Chi^2 = 23.47$, df = 7 (P = 0.001); $I^2 = 70\%$
Test for overall effect: Z = 3.87 (P = 0.0001)

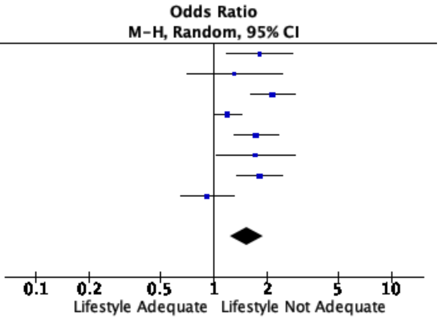


**Supplementary Figure 1.2B** Funnel Plot for Assessment of Publication Bias

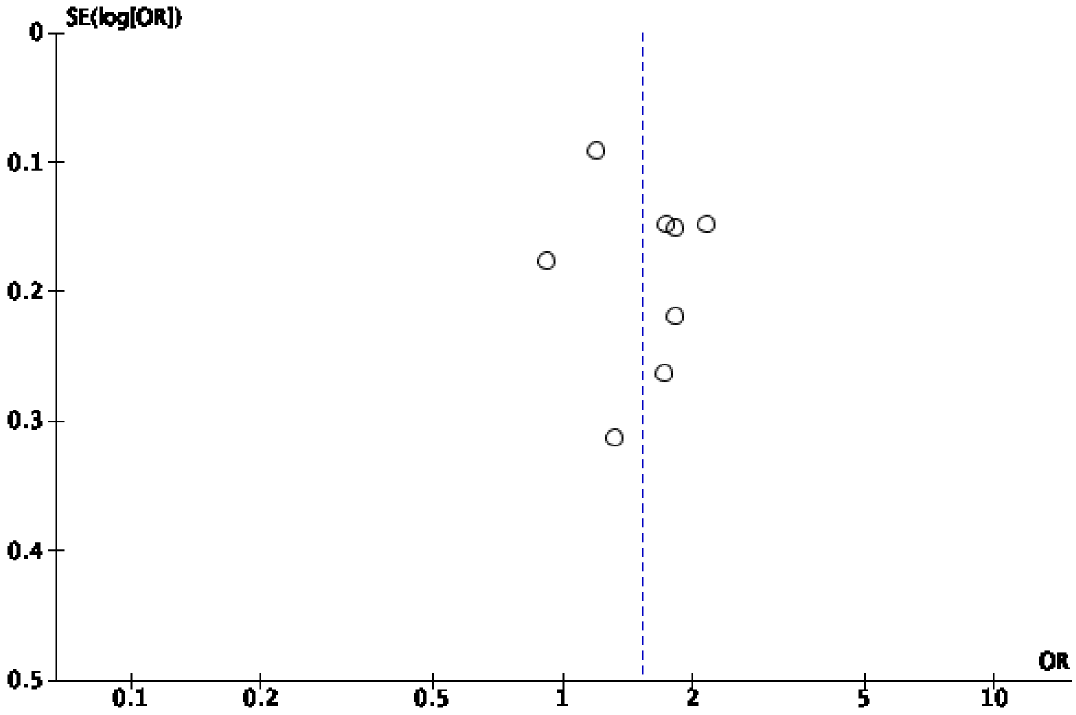

**Supplementary Figure 1.3A** Forest plot for included studies comparing if lifestyle was adequate or not adequate for body mass index

| | Lifestyle Adequate | | | Lifestyle Not Adequate | | | | Mean Difference |
|---|---|---|---|---|---|---|---|---|
| Study or Subgroup | Mean | SD | Total | Mean | SD | Total | Weight | IV, Random, 95% CI |
| Ares 2017 | 26.82 | 4.94 | 165 | 27.04 | 6.46 | 36 | 3.1% | −0.22 [−2.46, 2.02] |
| Barnes 2013 | 24.9 | 5.9 | 1171 | 28.62 | 6.76 | 524 | 8.2% | −3.72 [−4.39, −3.05] |
| Bashir 2020 | 31.2 | 6 | 358 | 32.9 | 6.3 | 443 | 7.5% | −1.70 [−2.55, −0.85] |
| Benhalima 2015 | 26.8 | 12.9 | 456 | 29.1 | 20.2 | 145 | 1.6% | −2.30 [−5.79, 1.19] |
| Ducarme 2019 | 27.4 | 5.8 | 126 | 28.6 | 6 | 72 | 4.4% | −1.20 [−2.92, 0.52] |
| Durnwald 2011 | 29.9 | 4.9 | 424 | 32.7 | 5.8 | 36 | 3.8% | −2.80 [−4.75, −0.85] |
| Ikenoue 2014 | 20.5 | 3.6 | 91 | 21.1 | 2.6 | 50 | 6.7% | −0.60 [−1.63, 0.43] |
| Ito 2016 | 23.3 | 3.6 | 70 | 24.6 | 4.3 | 32 | 4.4% | −1.30 [−3.01, 0.41] |
| Mecacci 2021 | 23.9 | 4.91 | 962 | 25.2 | 5.49 | 1012 | 9.0% | −1.30 [−1.76, −0.84] |
| Meghelli 2020 | 43.6 | 2.6 | 58 | 44 | 2.9 | 63 | 7.0% | −0.40 [−1.38, 0.58] |
| Ng 2020 | 29 | 7.42 | 1281 | 31.2 | 7.66 | 576 | 7.9% | −2.20 [−2.95, −1.45] |
| Nishikawa 2018 | 22.41 | 0.21 | 472 | 23.89 | 0.7 | 57 | 9.6% | −1.48 [−1.66, −1.30] |
| Souza 2019 | 29.8 | 6.9 | 273 | 32.2 | 6.4 | 135 | 5.5% | −2.40 [−3.75, −1.05] |
| Suhonen 2008 | 26.1 | 5.4 | 520 | 29 | 6.8 | 385 | 7.6% | −2.90 [−3.72, −2.08] |
| Sun 2021 | 23.9 | 4 | 616 | 24.8 | 4.3 | 92 | 7.1% | −0.90 [−1.83, 0.03] |
| Wong 2011 | 26.5 | 6.3 | 289 | 29.9 | 7.3 | 323 | 6.6% | −3.40 [−4.48, −2.32] |
| Total (95% CI) | | | 7332 | | | 3981 | 100.0% | −1.83 [−2.32, −1.35] |

Heterogeneity: $Tau^2 = 0.62$; $Chi^2 = 80.60$, $df = 15$ ($P < 0.00001$); $I^2 = 81\%$
Test for overall effect: $Z = 7.45$ ($P < 0.00001$)

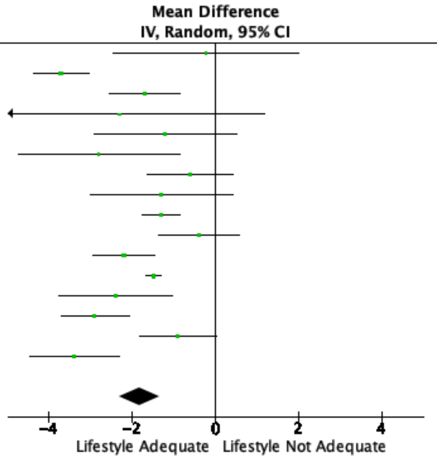


**Supplementary Figure 1.3B** Funnel Plot for Assessment of Publication Bias

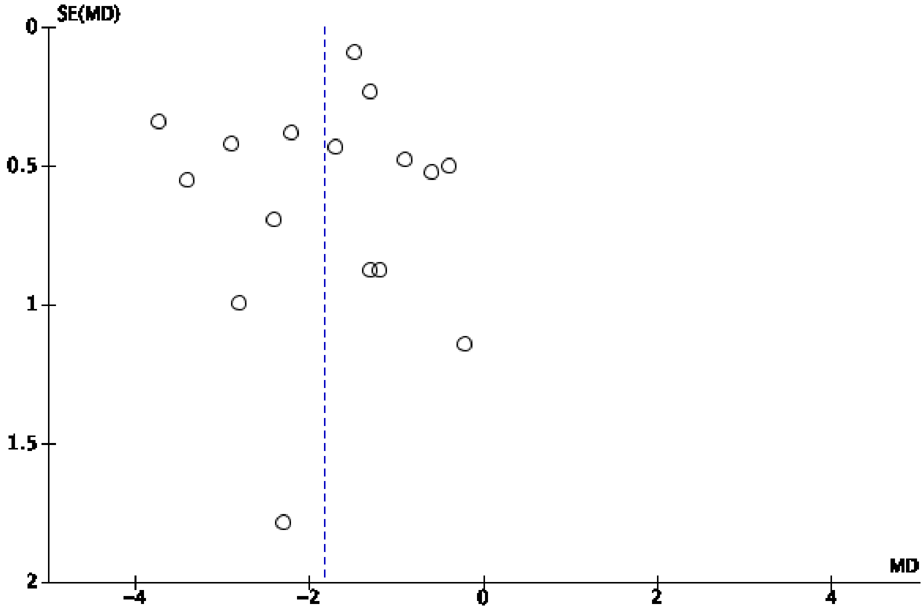

**Supplementary Figure 1.4A** Forest plot for included studies comparing if lifestyle was adequate or not adequate for previous history of gestational diabetes

| Study or Subgroup | Lifestyle Adequate Events | Lifestyle Adequate Total | Lifestyle Not Adequate Events | Lifestyle Not Adequate Total | Weight | Odds Ratio M-H, Random, 95% CI |
|---|---|---|---|---|---|---|
| Benhalima 2015 | 47 | 456 | 31 | 145 | 8.4% | 0.42 [0.26, 0.70] |
| Ducarme 2019 | 23 | 126 | 14 | 72 | 5.6% | 0.93 [0.44, 1.94] |
| Ikenoue 2014 | 1 | 91 | 0 | 50 | 0.5% | 1.67 [0.07, 41.86] |
| Kalok 2020 | 151 | 968 | 29 | 96 | 8.8% | 0.43 [0.27, 0.68] |
| Koning 2016 | 25 | 460 | 61 | 360 | 8.6% | 0.28 [0.17, 0.46] |
| Krispin 2021 | 61 | 314 | 76 | 328 | 10.2% | 0.80 [0.55, 1.17] |
| Mecacci 2021 | 85 | 962 | 151 | 1012 | 11.8% | 0.55 [0.42, 0.73] |
| Meghelli 2020 | 6 | 58 | 24 | 63 | 3.8% | 0.19 [0.07, 0.50] |
| Meshel 2016 | 293 | 1181 | 55 | 143 | 10.5% | 0.53 [0.37, 0.76] |
| Nishikawa 2018 | 25 | 472 | 5 | 57 | 3.7% | 0.58 [0.21, 1.58] |
| Ouzounian 2011 | 185 | 1189 | 89 | 262 | 11.5% | 0.36 [0.27, 0.48] |
| Souza 2019 | 17 | 273 | 32 | 135 | 6.7% | 0.21 [0.11, 0.40] |
| Wong 2011 | 48 | 289 | 84 | 323 | 9.9% | 0.57 [0.38, 0.84] |
| Total (95% CI) | | 6839 | | 3046 | 100.0% | 0.46 [0.37, 0.57] |
| Total events | 967 | | 651 | | | |

Heterogeneity: $Tau^2 = 0.09$; $Chi^2 = 31.08$, df = 12 (P = 0.002); $I^2 = 61\%$
Test for overall effect: Z = 6.86 (P < 0.00001)

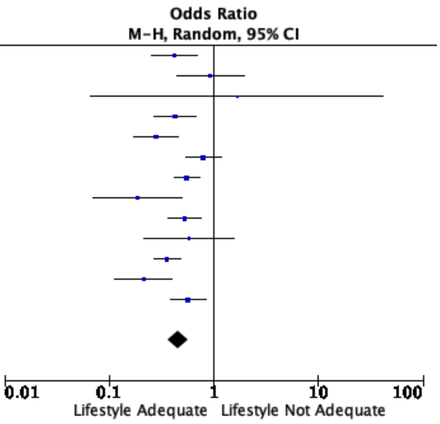


**Supplementary Figure 1.4B** Funnel Plot for Assessment of Publication Bias

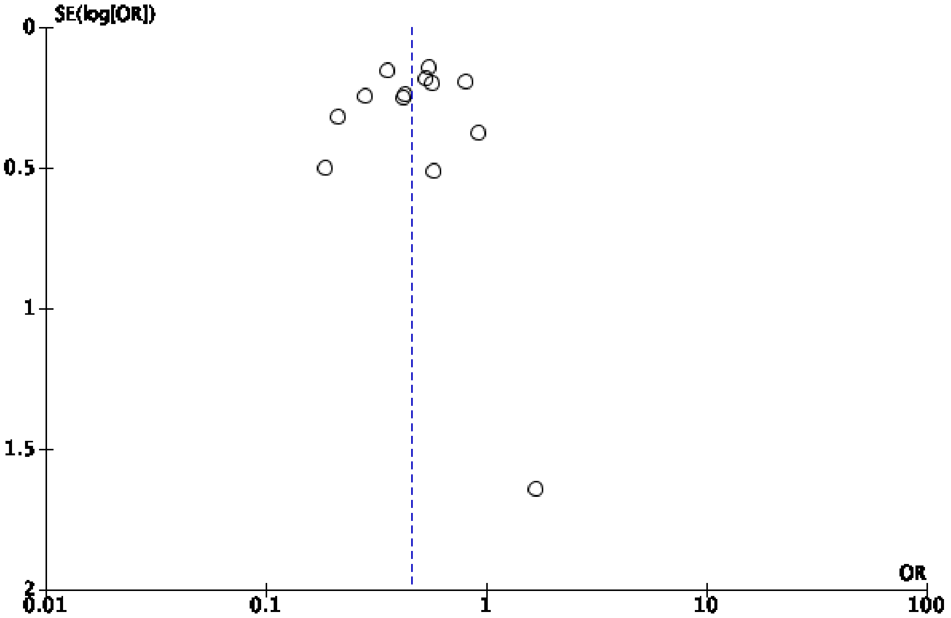

**Supplementary Figure 1.5A** Forest plot for included studies comparing if lifestyle was adequate or not adequate for haemoglobin A1C

| | Lifestyle Adequate | | | Lifestyle Not Adequate | | | | Mean Difference |
|---|---|---|---|---|---|---|---|---|
| Study or Subgroup | Mean | SD | Total | Mean | SD | Total | Weight | IV, Random, 95% CI |
| Barnes 2013 | 5.2 | 0.6 | 1171 | 5.5 | 0.6 | 524 | 14.3% | −0.30 [−0.36, −0.24] |
| Benhalima 2015 | 5.2 | 0.5 | 456 | 5.5 | 0.6 | 145 | 11.1% | −0.30 [−0.41, −0.19] |
| Ducarme 2019 | 5.2 | 0.3 | 126 | 5.3 | 0.4 | 72 | 11.3% | −0.10 [−0.21, 0.01] |
| Gilbert 2021 | 5.3 | 0.31 | 178 | 5.5 | 0.51 | 163 | 12.3% | −0.20 [−0.29, −0.11] |
| Ikenoue 2014 | 5.4 | 0.3 | 91 | 5.4 | 0.3 | 50 | 11.4% | 0.00 [−0.10, 0.10] |
| Nishikawa 2018 | 5.24 | 0.01 | 472 | 5.46 | 0.06 | 57 | 16.5% | −0.22 [−0.24, −0.20] |
| Sun 2021 | 5.2 | 0.4 | 616 | 5.6 | 0.6 | 92 | 9.9% | −0.40 [−0.53, −0.27] |
| Wong 2011 | 5.41 | 0.39 | 289 | 5.54 | 0.61 | 323 | 13.1% | −0.13 [−0.21, −0.05] |
| Total (95% CI) | | | 3399 | | | 1426 | 100.0% | −0.21 [−0.27, −0.14] |

Heterogeneity: $Tau^2 = 0.01$; $Chi^2 = 43.53$, df = 7 ($P < 0.00001$); $I^2 = 84\%$
Test for overall effect: Z = 6.44 ($P < 0.00001$)

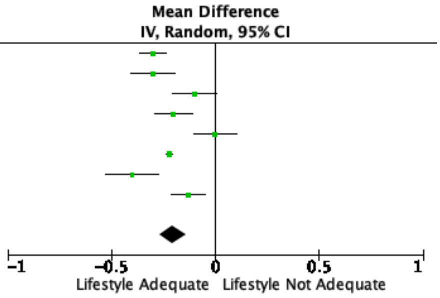


**Supplementary Figure 1.5B** Funnel Plot for Assessment of Publication Bias

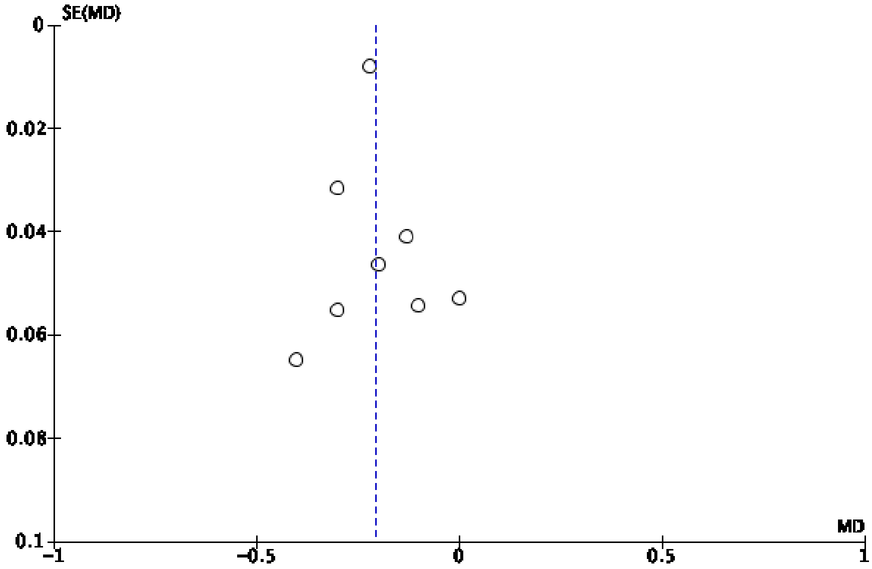

**Supplementary Figure 1.6A** Forest plot for included studies comparing if lifestyle was adequate or not adequate for fasting glucose

| | Lifestyle Adequate | | | Lifestyle Not Adequate | | | | Mean Difference |
|---|---|---|---|---|---|---|---|---|
| Study or Subgroup | Mean | SD | Total | Mean | SD | Total | Weight | IV, Random, 95% CI |
| Ares 2017 | 87.9 | 9.02 | 165 | 95.68 | 11.26 | 36 | 6.8% | −7.78 [−11.71, −3.85] |
| Bashir 2020 | 86.4 | 9 | 358 | 97.2 | 12.6 | 443 | 8.4% | −10.80 [−12.30, −9.30] |
| Benhalima 2015 | 87.7 | 10.3 | 456 | 97.6 | 18.8 | 145 | 7.3% | −9.90 [−13.10, −6.70] |
| Ducarme 2019 | 92 | 8 | 126 | 95 | 12 | 72 | 7.4% | −3.00 [−6.10, 0.10] |
| Ikenoue 2014 | 88.9 | 7.4 | 91 | 89.3 | 7.1 | 50 | 7.8% | −0.40 [−2.89, 2.09] |
| Ito 2016 | 89.6 | 8.7 | 70 | 91.5 | 7.9 | 32 | 7.2% | −1.90 [−5.31, 1.51] |
| Kalok 2020 | 84.78 | 10.8 | 968 | 94.5 | 13.32 | 96 | 7.6% | −9.72 [−12.47, −6.97] |
| Mecacci 2021 | 86.71 | 23.39 | 962 | 90.07 | 9.7 | 1012 | 8.3% | −3.36 [−4.95, −1.77] |
| Meshel 2016 | 88.09 | 11.91 | 1181 | 100.31 | 10.86 | 143 | 8.2% | −12.22 [−14.13, −10.31] |
| Nishikawa 2018 | 91.5 | 0.4 | 472 | 94.1 | 1.4 | 57 | 8.7% | −2.60 [−2.97, −2.23] |
| Souza 2019 | 96.8 | 5.2 | 273 | 101.1 | 7.8 | 135 | 8.4% | −4.30 [−5.75, −2.85] |
| Sun 2021 | 84.6 | 13.14 | 616 | 100.98 | 26.64 | 92 | 5.6% | −16.38 [−21.92, −10.84] |
| Wong 2011 | 97.38 | 7.02 | 289 | 99.72 | 10.98 | 323 | 8.4% | −2.34 [−3.79, −0.89] |
| Total (95% CI) | | | 6027 | | | 2636 | 100.0% | −6.26 [−8.44, −4.08] |

Heterogeneity: $Tau^2 = 14.18$; $Chi^2 = 265.84$, df = 12 ($P < 0.00001$); $I^2 = 95\%$
Test for overall effect: $Z = 5.63$ ($P < 0.00001$)

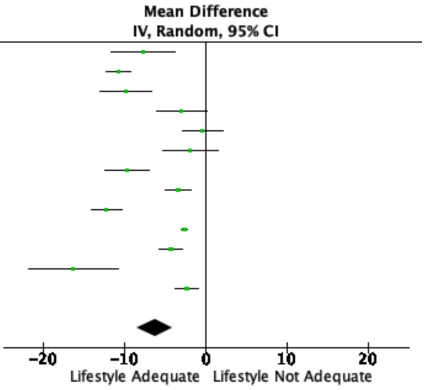


**Supplementary Figure 1.6B** Funnel Plot for Assessment of Publication Bias

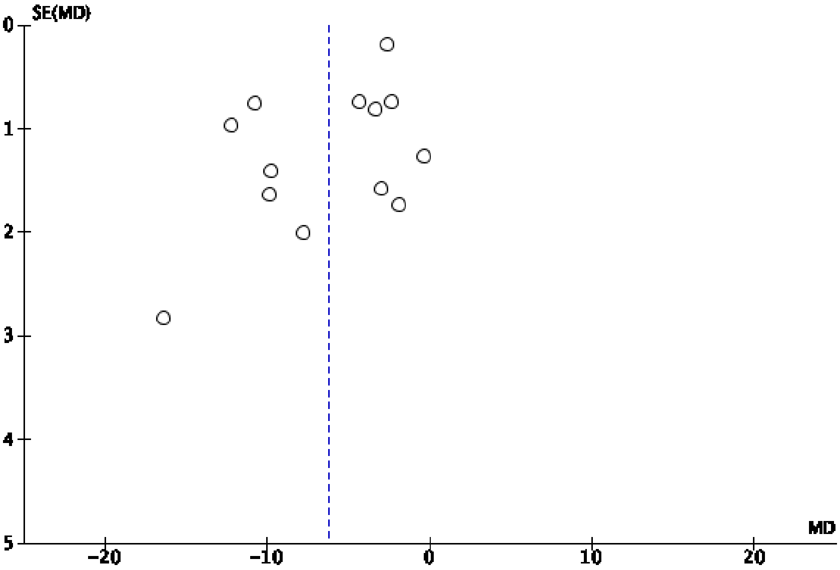

**Supplementary Figure 1.7A** Forest plot for included studies comparing if lifestyle was adequate or not adequate for 1-hour glucose

| | Lifestyle Adequate | | | Lifestyle Not Adequate | | | | Mean Difference |
|---|---|---|---|---|---|---|---|---|
| Study or Subgroup | Mean | SD | Total | Mean | SD | Total | Weight | IV, Random, 95% CI |
| Ares 2017 | 195.16 | 28 | 165 | 202 | 42.85 | 36 | 6.4% | −6.84 [−21.47, 7.79] |
| Bashir 2020 | 178.2 | 30.6 | 358 | 196.2 | 28.8 | 443 | 11.2% | −18.00 [−22.15, −13.85] |
| Benhalima 2015 | 184.5 | 25.8 | 456 | 194.7 | 30.1 | 145 | 10.7% | −10.20 [−15.64, −4.76] |
| Ducarme 2019 | 167 | 0.26 | 126 | 182 | 0.31 | 72 | 11.9% | −15.00 [−15.08, −14.92] |
| Ikenoue 2014 | 153.7 | 27.3 | 91 | 152.8 | 29 | 50 | 8.6% | 0.90 [−8.90, 10.70] |
| Ito 2016 | 155.3 | 33.6 | 70 | 179.9 | 34.9 | 32 | 6.5% | −24.60 [−39.03, −10.17] |
| Mecacci 2021 | 171.48 | 21.12 | 962 | 177.71 | 21.16 | 1012 | 11.8% | −6.23 [−8.10, −4.36] |
| Meshel 2016 | 88.09 | 11.91 | 1181 | 100.31 | 10.86 | 143 | 11.7% | −12.22 [−14.13, −10.31] |
| Nishikawa 2018 | 155.4 | 1.5 | 472 | 184.9 | 4.4 | 57 | 11.9% | −29.50 [−30.65, −28.35] |
| Sun 2021 | 173.52 | 29.34 | 616 | 203.76 | 39.6 | 92 | 9.3% | −30.24 [−38.66, −21.82] |
| Total (95% CI) | | | 4497 | | | 2082 | 100.0% | −15.33 [−20.81, −9.85] |

Heterogeneity: $Tau^2 = 65.65$; $Chi^2 = 732.47$, $df = 9$ ($P < 0.00001$); $I^2 = 99\%$
Test for overall effect: $Z = 5.48$ ($P < 0.00001$)

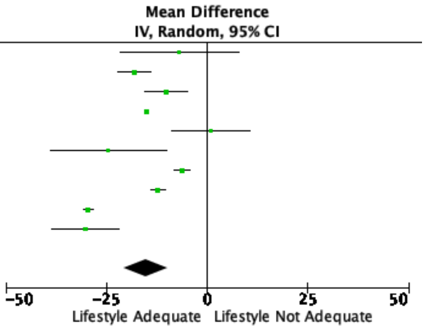


**Supplementary Figure 1.7B** Funnel Plot for Assessment of Publication Bias

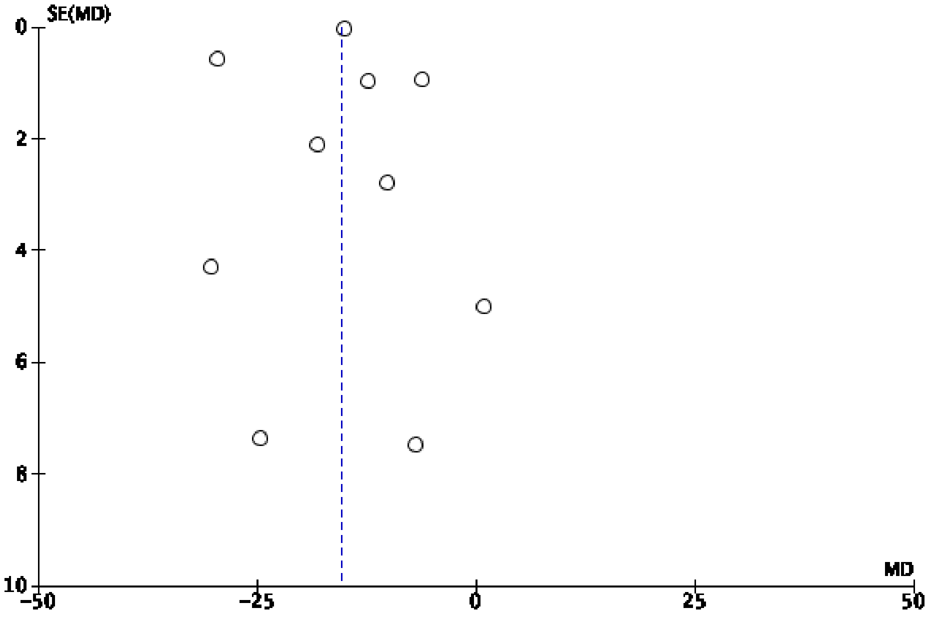

**Supplementary Figure 1.8A** Forest plot for included studies comparing if lifestyle was adequate or not adequate for 2-hour glucose

| | Lifestyle Adequate | | | Lifestyle Not Adequate | | | | Mean Difference |
|---|---|---|---|---|---|---|---|---|
| Study or Subgroup | Mean | SD | Total | Mean | SD | Total | Weight | IV, Random, 95% CI |
| Ares 2017 | 179.53 | 22.4 | 165 | 190.78 | 47.33 | 36 | 4.5% | −11.25 [−27.08, 4.58] |
| Bashir 2020 | 149.4 | 28.8 | 358 | 165.6 | 84.6 | 443 | 7.6% | −16.20 [−24.62, −7.78] |
| Benhalima 2015 | 175 | 22.8 | 456 | 185.2 | 28.5 | 145 | 9.2% | −10.20 [−15.29, −5.11] |
| Ducarme 2019 | 151 | 0.25 | 126 | 157 | 0.32 | 72 | 10.4% | −6.00 [−6.09, −5.91] |
| Ikenoue 2014 | 135.8 | 26.2 | 91 | 146.6 | 29.6 | 50 | 6.9% | −10.80 [−20.61, −0.99] |
| Ito 2016 | 135.3 | 29.7 | 70 | 150.5 | 28.5 | 32 | 5.9% | −15.20 [−27.28, −3.12] |
| Kalok 2020 | 152.82 | 14.58 | 968 | 156.24 | 18 | 96 | 9.7% | −3.42 [−7.14, 0.30] |
| Mecacci 2021 | 138.06 | 30.08 | 962 | 140.45 | 33.29 | 1012 | 10.0% | −2.39 [−5.19, 0.41] |
| Meshel 2016 | 162.22 | 29.68 | 1181 | 161.69 | 36.31 | 143 | 8.7% | 0.53 [−5.66, 6.72] |
| Nishikawa 2018 | 139.5 | 1.2 | 472 | 157.1 | 4.1 | 57 | 10.3% | −17.60 [−18.67, −16.53] |
| Sun 2021 | 149.22 | 27.9 | 616 | 172.98 | 44.82 | 92 | 7.1% | −23.76 [−33.18, −14.34] |
| Wong 2011 | 149.4 | 22.68 | 289 | 150.12 | 26.64 | 323 | 9.6% | −0.72 [−4.63, 3.19] |
| Total (95% CI) | | | 5754 | | | 2501 | 100.0% | −9.06 [−13.55, −4.56] |

Heterogeneity: $Tau^2 = 50.61$; $Chi^2 = 494.03$, df = 11 ($P < 0.00001$); $I^2 = 98\%$
Test for overall effect: Z = 3.95 ($P < 0.0001$)

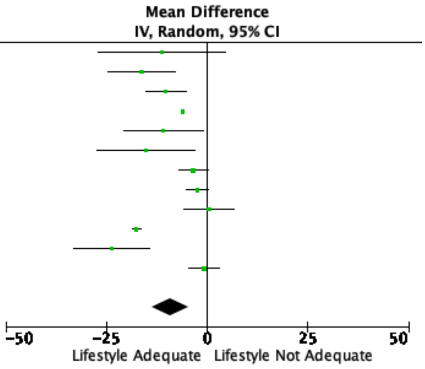


**Supplementary Figure 1.8B** Funnel Plot for Assessment of Publication Bias

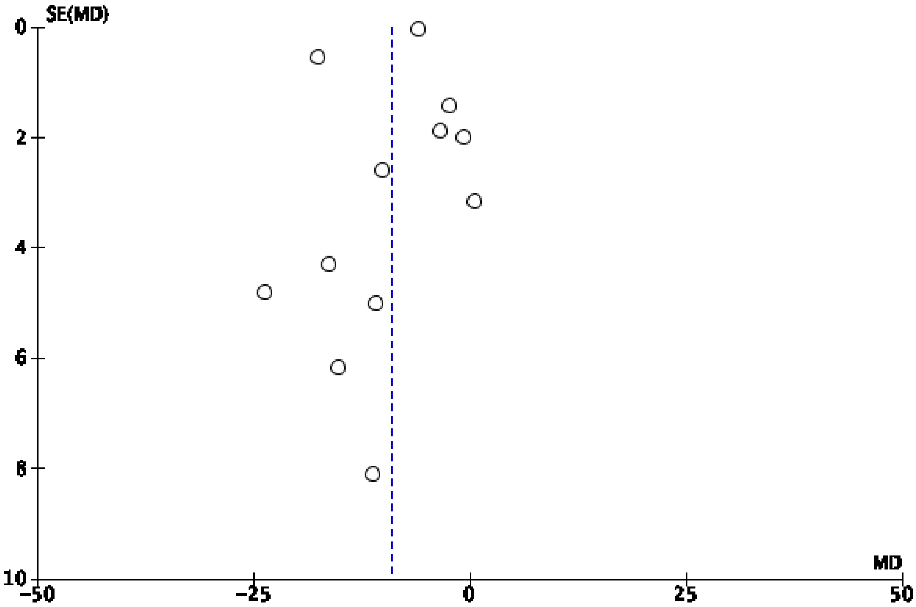

**Supplementary Figure 1.9A** Forest plot for included studies comparing if lifestyle was adequate or not adequate for 3-hour glucose

| | Lifestyle Adequate | | | Lifestyle Not Adequate | | | | Mean Difference |
|---|---|---|---|---|---|---|---|---|
| Study or Subgroup | Mean | SD | Total | Mean | SD | Total | Weight | IV, Random, 95% CI |
| Ares 2017 | 152.77 | 25.72 | 165 | 161.47 | 31.52 | 36 | 13.3% | −8.70 [−19.72, 2.32] |
| Benhalima 2015 | 145.4 | 27.6 | 456 | 152.7 | 33.4 | 145 | 45.0% | −7.30 [−13.30, −1.30] |
| Meshel 2016 | 97.87 | 35.49 | 1181 | 107.74 | 35.94 | 143 | 41.7% | −9.87 [−16.10, −3.64] |
| Total (95% CI) | | | 1802 | | | 324 | 100.0% | −8.56 [−12.58, −4.54] |

Heterogeneity: Tau² = 0.00; Chi² = 0.34, df = 2 (P = 0.84); I² = 0%
Test for overall effect: Z = 4.17 (P < 0.0001)

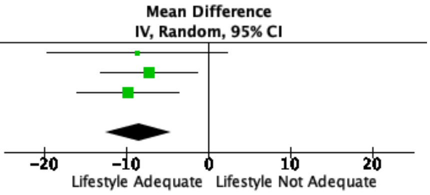


**Supplementary Figure 1.9B** Funnel Plot for Assessment of Publication Bias

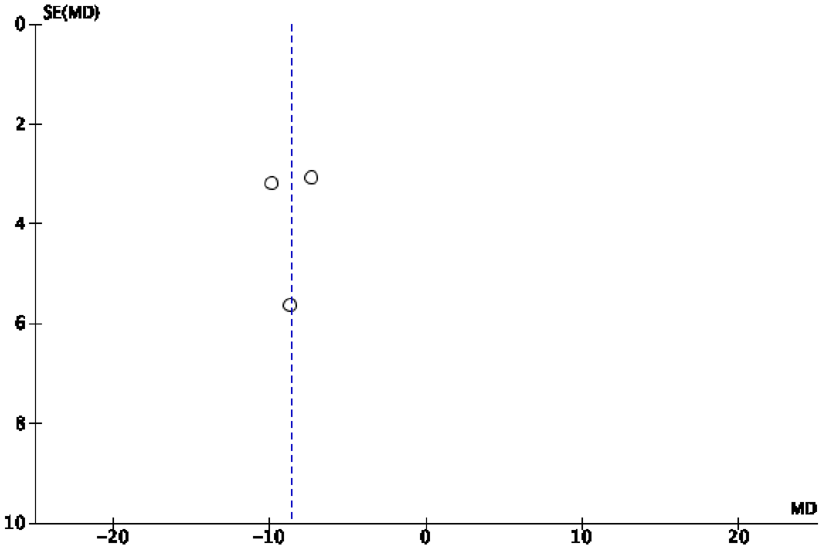

**Supplementary Figure 1.10A** Forest plot for included studies comparing if lifestyle was adequate or not adequate for family history of diabetes

| Study or Subgroup | Lifestyle Adequate Events | Lifestyle Adequate Total | Lifestyle Not Adequate Events | Lifestyle Not Adequate Total | Weight | Odds Ratio M-H, Random, 95% CI |
|---|---|---|---|---|---|---|
| Benhalima 2015 | 75 | 456 | 33 | 135 | 6.1% | 0.61 [0.38, 0.97] |
| Gilbert 2021 | 97 | 178 | 96 | 163 | 6.9% | 0.84 [0.54, 1.28] |
| Ikenoue 2014 | 12 | 91 | 10 | 50 | 1.7% | 0.61 [0.24, 1.53] |
| Ito 2016 | 16 | 70 | 15 | 32 | 1.9% | 0.34 [0.14, 0.82] |
| Kalok 2020 | 210 | 968 | 30 | 96 | 6.2% | 0.61 [0.39, 0.96] |
| Koning 2016 | 156 | 460 | 170 | 360 | 13.0% | 0.57 [0.43, 0.76] |
| Krispin 2021 | 95 | 314 | 123 | 328 | 10.5% | 0.72 [0.52, 1.00] |
| Mecacci 2021 | 223 | 962 | 309 | 1012 | 19.6% | 0.69 [0.56, 0.84] |
| Meshel 2016 | 479 | 1181 | 66 | 143 | 9.6% | 0.80 [0.56, 1.13] |
| Nishikawa 2018 | 199 | 472 | 38 | 57 | 4.1% | 0.36 [0.20, 0.65] |
| Souza 2019 | 159 | 273 | 98 | 135 | 6.5% | 0.53 [0.34, 0.82] |
| Sun 2021 | 46 | 616 | 10 | 92 | 2.8% | 0.66 [0.32, 1.36] |
| Wong 2011 | 134 | 289 | 157 | 323 | 11.1% | 0.91 [0.67, 1.26] |
| Total (95% CI) | | 6330 | | 2926 | 100.0% | 0.66 [0.59, 0.75] |
| Total events | 1901 | | 1155 | | | |

Heterogeneity: $Tau^2 = 0.01$; $Chi^2 = 15.07$, df = 12 (P = 0.24); $I^2 = 20\%$
Test for overall effect: Z = 6.45 (P < 0.00001)

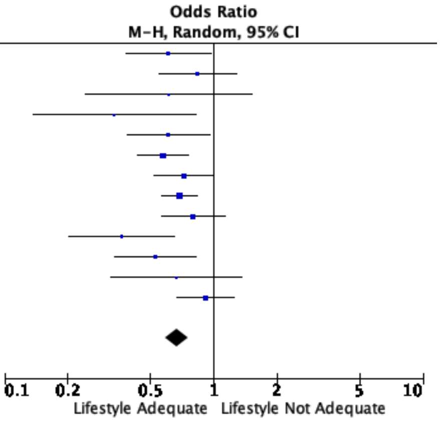


**Supplementary Figure 1.10B** Funnel Plot for Assessment of Publication Bias

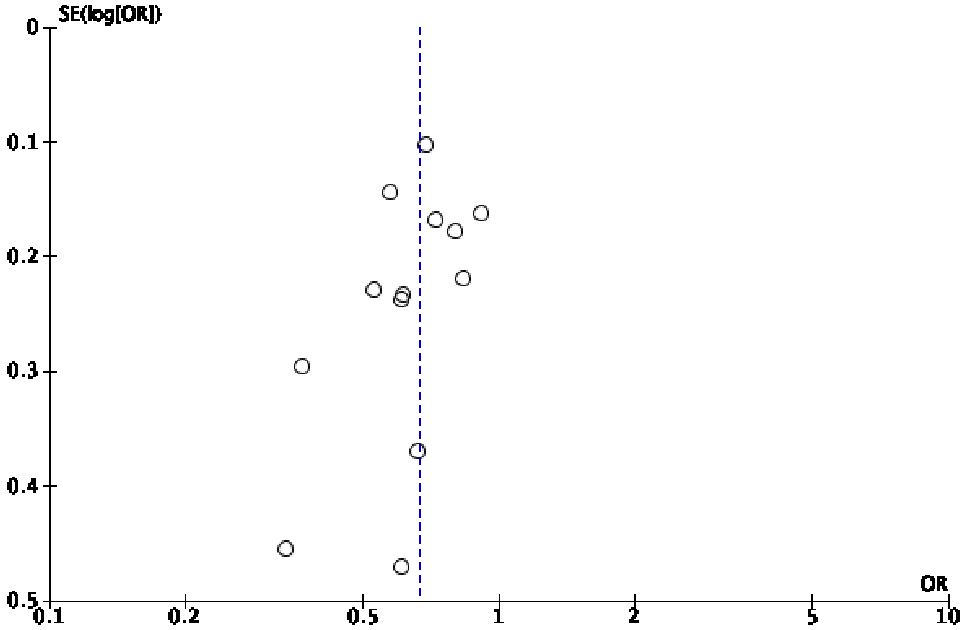

**Supplementary Figure 1.11A** Forest plot for included studies comparing if lifestyle was adequate or not adequate for gestational age at gestational diabetes diagnosis

| Study or Subgroup | Lifestyle Adequate Events | Lifestyle Adequate Total | Lifestyle Not Adequate Events | Lifestyle Not Adequate Total | Weight | Odds Ratio M-H, Random, 95% CI |
|---|---|---|---|---|---|---|
| Benhalima 2015 | 75 | 456 | 33 | 135 | 6.1% | 0.61 [0.38, 0.97] |
| Gilbert 2021 | 97 | 178 | 96 | 163 | 6.9% | 0.84 [0.54, 1.28] |
| Ikenoue 2014 | 12 | 91 | 10 | 50 | 1.7% | 0.61 [0.24, 1.53] |
| Ito 2016 | 16 | 70 | 15 | 32 | 1.9% | 0.34 [0.14, 0.82] |
| Kalok 2020 | 210 | 968 | 30 | 96 | 6.2% | 0.61 [0.39, 0.96] |
| Koning 2016 | 156 | 460 | 170 | 360 | 13.0% | 0.57 [0.43, 0.76] |
| Krispin 2021 | 95 | 314 | 123 | 328 | 10.5% | 0.72 [0.52, 1.00] |
| Mecacci 2021 | 223 | 962 | 309 | 1012 | 19.6% | 0.69 [0.56, 0.84] |
| Meshel 2016 | 479 | 1181 | 66 | 143 | 9.6% | 0.80 [0.56, 1.13] |
| Nishikawa 2018 | 199 | 472 | 38 | 57 | 4.1% | 0.36 [0.20, 0.65] |
| Souza 2019 | 159 | 273 | 98 | 135 | 6.5% | 0.53 [0.34, 0.82] |
| Sun 2021 | 46 | 616 | 10 | 92 | 2.8% | 0.66 [0.32, 1.36] |
| Wong 2011 | 134 | 289 | 157 | 323 | 11.1% | 0.91 [0.67, 1.26] |
| Total (95% CI) | | 6330 | | 2926 | 100.0% | 0.66 [0.59, 0.75] |
| Total events | 1901 | | 1155 | | | |

Heterogeneity: $Tau^2 = 0.01$; $Chi^2 = 15.07$, df = 12 (P = 0.24); $I^2 = 20\%$
Test for overall effect: Z = 6.45 (P < 0.00001)

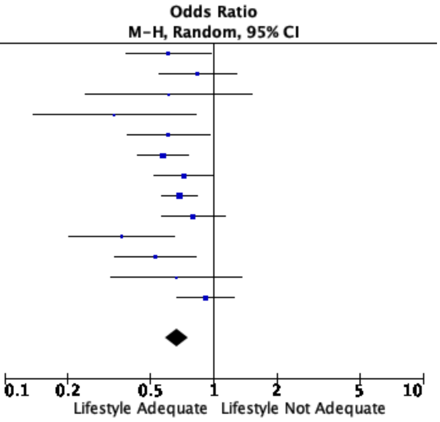


**Supplementary Figure 1.11B** Funnel Plot for Assessment of Publication Bias

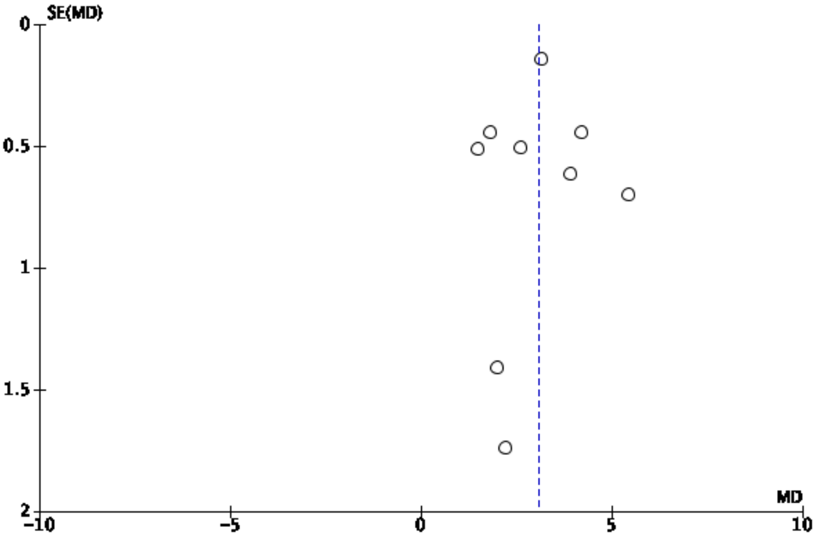

**Supplementary Figure 1.12A** Forest plot for included studies comparing if lifestyle was adequate or not adequate for history of smoking

| Study or Subgroup | Lifestyle Adequate Events | Lifestyle Adequate Total | Lifestyle Not Adequate Events | Lifestyle Not Adequate Total | Weight | Odds Ratio M-H, Random, 95% CI |
|---|---|---|---|---|---|---|
| Durnwald 2011 | 30 | 424 | 6 | 36 | 13.1% | 0.38 [0.15, 0.99] |
| Krispin 2021 | 22 | 314 | 29 | 328 | 22.3% | 0.78 [0.44, 1.38] |
| Meghelli 2020 | 10 | 58 | 12 | 63 | 13.5% | 0.89 [0.35, 2.24] |
| Ng 2020 | 218 | 1281 | 81 | 576 | 32.6% | 1.25 [0.95, 1.65] |
| Souza 2019 | 19 | 273 | 15 | 135 | 18.4% | 0.60 [0.29, 1.22] |
| Total (95% CI) | | 2350 | | 1138 | 100.0% | 0.80 [0.52, 1.23] |
| Total events | 299 | | 143 | | | |

Heterogeneity: $Tau^2 = 0.12$; $Chi^2 = 9.35$, df = 4 (P = 0.05); $I^2 = 57\%$
Test for overall effect: Z = 1.01 (P = 0.31)

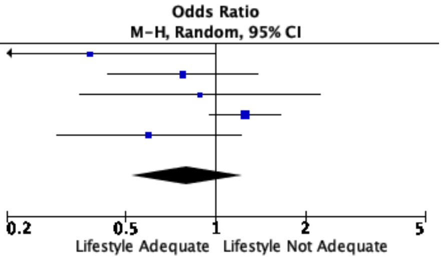


**Supplementary Figure 1.12B** Funnel Plot for Assessment of Publication Bias

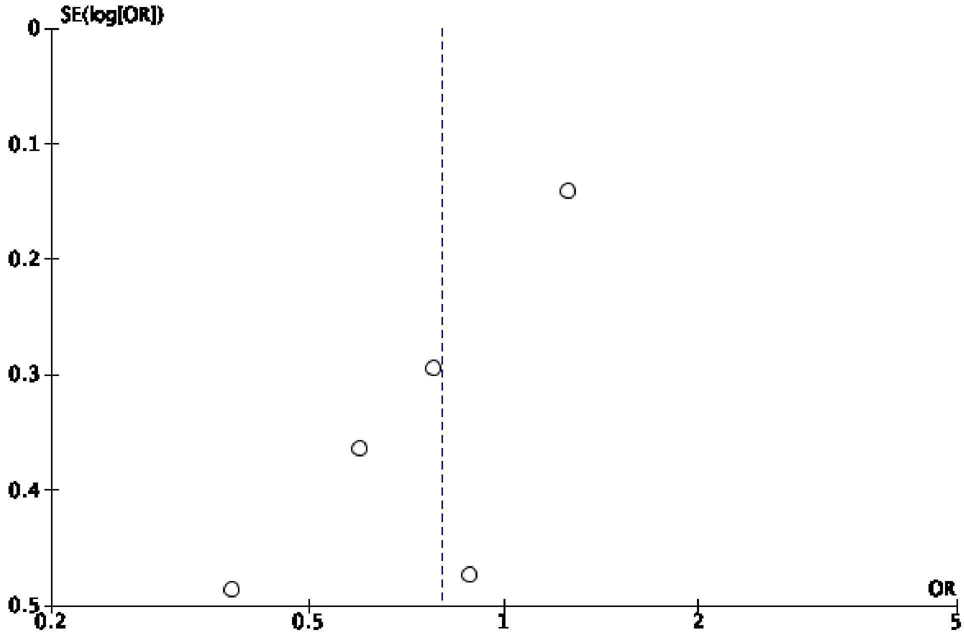

**Supplementary Figure 1.13A** Forest plot for included studies comparing if lifestyle was adequate or not adequate for previous history of macrosomia

| Study or Subgroup | Lifestyle Adequate Events | Lifestyle Adequate Total | Lifestyle Not Adequate Events | Lifestyle Not Adequate Total | Weight | Odds Ratio M-H, Random, 95% CI |
|---|---|---|---|---|---|---|
| Ducarme 2019 | 7 | 126 | 8 | 72 | 9.7% | 0.47 [0.16, 1.36] |
| Ito 2016 | 0 | 70 | 1 | 32 | 1.5% | 0.15 [0.01, 3.76] |
| Koning 2016 | 55 | 460 | 35 | 360 | 20.6% | 1.26 [0.81, 1.97] |
| Krispin 2021 | 19 | 314 | 29 | 328 | 17.2% | 0.66 [0.36, 1.21] |
| Mecacci 2021 | 11 | 962 | 14 | 1012 | 13.5% | 0.82 [0.37, 1.83] |
| Ouzounian 2011 | 116 | 1189 | 49 | 262 | 22.4% | 0.47 [0.33, 0.68] |
| Souza 2019 | 16 | 273 | 19 | 135 | 15.2% | 0.38 [0.19, 0.77] |
| Total (95% CI) | | 3394 | | 2201 | 100.0% | 0.63 [0.42, 0.94] |
| Total events | 224 | | 155 | | | |

Heterogeneity: $Tau^2 = 0.16$; $Chi^2 = 15.23$, df = 6 (P = 0.02); $I^2 = 61\%$
Test for overall effect: Z = 2.23 (P = 0.03)

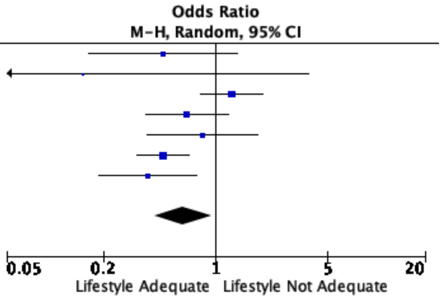


**Supplementary Figure 1.13B** Funnel Plot for Assessment of Publication Bias

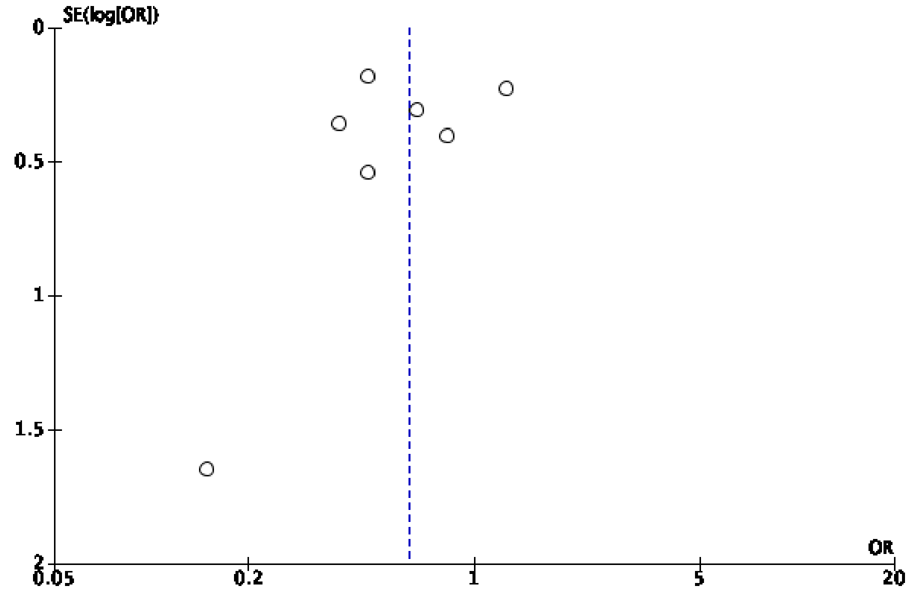

**Supplementary Figures 2.1 to 2.12**

**Forest Plots (A) and Funnel Plots (B) for Oral Pharmacological Agent adequate in controlling glucose vs not adequate**

**Supplementary Figure 2.1A** Forest plot for included studies comparing if oral pharmacological agent was adequate or not adequate for maternal age

| | Oral agent success | | | Oral agent failure | | | | Mean Difference |
|---|---|---|---|---|---|---|---|---|
| Study or Subgroup | Mean | SD | Total | Mean | SD | Total | Weight | IV, Random, 95% CI |
| Chmait 2004 | 32 | 5 | 56 | 31 | 7 | 13 | 4.8% | 1.00 [−3.02, 5.02] |
| Conway 2004 | 31.3 | 6.4 | 63 | 30.3 | 5 | 12 | 6.4% | 1.00 [−2.24, 4.24] |
| Gante 2018 | 32.9 | 5.4 | 253 | 35.2 | 5 | 135 | 13.6% | −2.30 [−3.37, −1.23] |
| Harper 2016 | 29.7 | 5.8 | 157 | 30.1 | 5.1 | 63 | 11.8% | −0.40 [−1.95, 1.15] |
| Kahn 2006 | 29 | 5 | 77 | 34 | 5 | 18 | 8.2% | −5.00 [−7.57, −2.43] |
| Khin 2018 | 30 | 5.9 | 61 | 32.4 | 5.5 | 77 | 10.4% | −2.40 [−4.32, −0.48] |
| McGrath 2016 | 32.9 | 4.6 | 29 | 32.8 | 5.3 | 34 | 8.6% | 0.10 [−2.34, 2.54] |
| Picón-César 2021 | 34.84 | 5.75 | 70 | 33.75 | 3.85 | 20 | 9.6% | 1.09 [−1.07, 3.25] |
| Rochon 2006 | 30.5 | 5.8 | 80 | 31.3 | 6.2 | 21 | 7.2% | −0.80 [−3.74, 2.14] |
| Tertti 2013 | 31.4 | 4 | 87 | 33.8 | 4.9 | 23 | 9.5% | −2.40 [−4.57, −0.23] |
| Yogev 2011 | 33.2 | 5 | 93 | 32.7 | 5.3 | 31 | 9.7% | 0.50 [−1.62, 2.62] |
| Total (95% CI) | | | 1026 | | | 447 | 100.0% | −1.04 [−2.10, 0.03] |

Heterogeneity: $Tau^2 = 1.86$; $Chi^2 = 26.88$, df = 10 ($P = 0.003$); $I^2 = 63\%$
Test for overall effect: $Z = 1.91$ ($P = 0.06$)

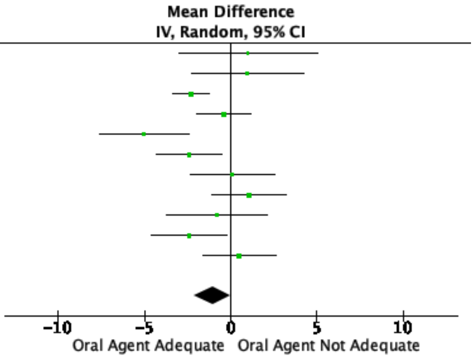


**Supplementary Figure 2.1B** Funnel Plot for Assessment of Publication Bias

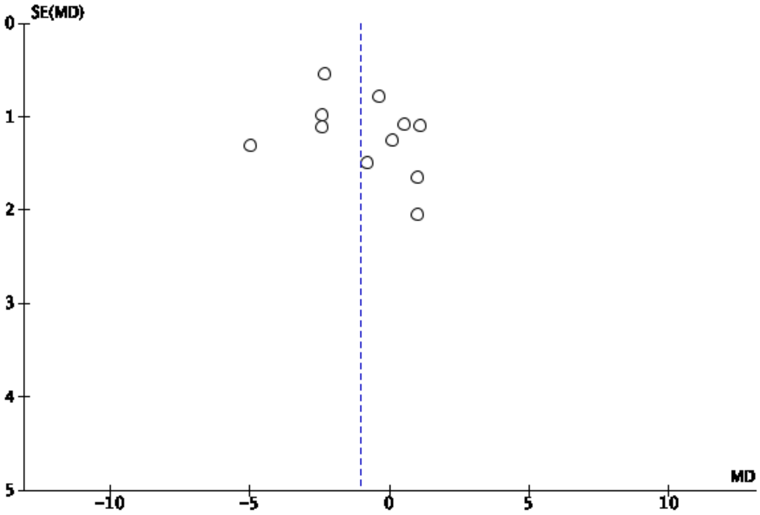

**Supplementary Figure 2.2A** Forest plot for included studies comparing if oral pharmacological agent was adequate or not adequate for nulliparity

| Study or Subgroup | Oral agent success Events | Oral agent success Total | Oral agent failure Events | Oral agent failure Total | Weight | Odds Ratio M-H, Random, 95% CI |
|---|---|---|---|---|---|---|
| Chmait 2004 | 8 | 56 | 1 | 13 | 1.6% | 2.00 [0.23, 17.57] |
| Harper 2016 | 47 | 157 | 14 | 63 | 16.5% | 1.50 [0.75, 2.97] |
| Khin 2018 | 21 | 61 | 23 | 77 | 14.9% | 1.23 [0.60, 2.53] |
| Picón-César 2021 | 44 | 70 | 12 | 20 | 7.5% | 1.13 [0.41, 3.12] |
| Rochon 2006 | 24 | 80 | 3 | 21 | 4.5% | 2.57 [0.69, 9.55] |
| Rowan 2008 | 75 | 195 | 40 | 168 | 36.9% | 2.00 [1.27, 3.16] |
| Tertti 2013 | 34 | 87 | 8 | 23 | 8.4% | 1.20 [0.46, 3.14] |
| Yogev 2011 | 28 | 93 | 9 | 31 | 9.7% | 1.05 [0.43, 2.57] |
| Total (95% CI) | | 799 | | 416 | 100.0% | 1.55 [1.17, 2.04] |
| Total events | 281 | | 110 | | | |

Heterogeneity: $Tau^2 = 0.00$; $Chi^2 = 3.58$, $df = 7$ ($P = 0.83$); $I^2 = 0\%$
Test for overall effect: $Z = 3.08$ ($P = 0.002$)

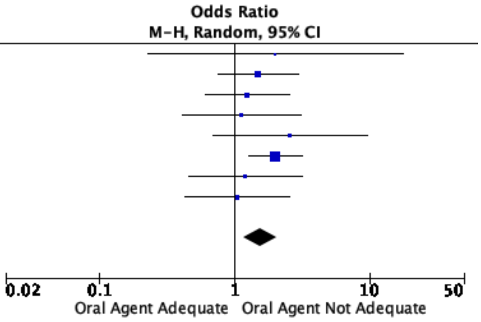


**Supplementary Figure 2.2B** Funnel Plot for Assessment of Publication Bias

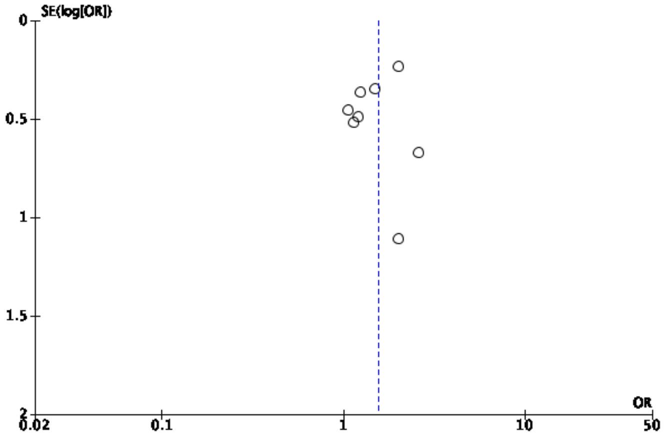

**Supplementary Figure 2.3A** Forest plot for included studies comparing if oral pharmacological agent was adequate or not adequate for body mass index

| Study or Subgroup | Oral agent success Mean | SD | Total | Oral agent failure Mean | SD | Total | Weight | Mean Difference IV, Random, 95% CI |
|---|---|---|---|---|---|---|---|---|
| Gante 2018 | 29 | 6.3 | 253 | 31.8 | 6.6 | 135 | 17.0% | −2.80 [−4.16, −1.44] |
| Harper 2016 | 35 | 8.6 | 157 | 35.4 | 8.2 | 63 | 10.0% | −0.40 [−2.83, 2.03] |
| Kahn 2006 | 30 | 6 | 77 | 32 | 8 | 18 | 5.1% | −2.00 [−5.93, 1.93] |
| Khin 2018 | 32.4 | 7.1 | 61 | 32.7 | 7.1 | 77 | 10.3% | −0.30 [−2.69, 2.09] |
| McGrath 2016 | 25.9 | 8.3 | 29 | 30 | 7.8 | 34 | 5.0% | −4.10 [−8.10, −0.10] |
| Picón-César 2021 | 29.75 | 6.26 | 70 | 30.65 | 4.58 | 20 | 9.8% | −0.90 [−3.39, 1.59] |
| Rochon 2006 | 31.5 | 7.5 | 80 | 32.2 | 5 | 21 | 8.8% | −0.70 [−3.40, 2.00] |
| Rowan 2008 | 31.1 | 7.8 | 195 | 33.6 | 8.6 | 168 | 14.4% | −2.50 [−4.20, −0.80] |
| Tertti 2013 | 29.8 | 6 | 87 | 28.2 | 5.8 | 23 | 8.9% | 1.60 [−1.08, 4.28] |
| Yogev 2011 | 28 | 5.4 | 93 | 27.9 | 5.8 | 31 | 10.6% | 0.10 [−2.22, 2.42] |
| **Total (95% CI)** | | | 1102 | | | 590 | 100.0% | −1.21 [−2.21, −0.21] |

Heterogeneity: $Tau^2 = 1.05$; $Chi^2 = 15.80$, df = 9 (P = 0.07); $I^2 = 43\%$
Test for overall effect: Z = 2.38 (P = 0.02)

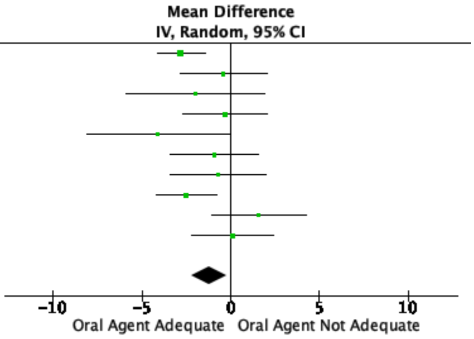


**Supplementary Figure 2.3B** Funnel Plot for Assessment of Publication Bias

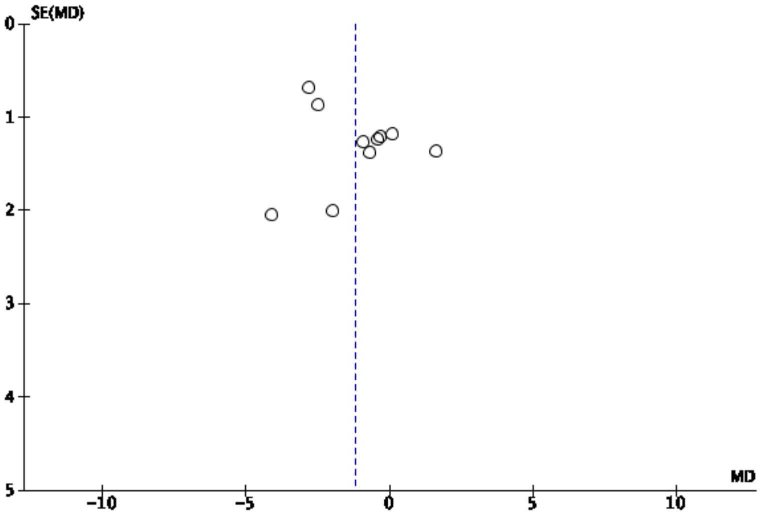

**Supplementary Figure 2.4A** Forest plot for included studies comparing if oral pharmacological agent was adequate or not adequate for previous history of gestational diabetes

| Study or Subgroup | Oral agent success Events | Oral agent success Total | Oral agent failure Events | Oral agent failure Total | Weight | Odds Ratio M-H, Random, 95% CI |
|---|---|---|---|---|---|---|
| Chmait 2004 | 9 | 56 | 4 | 13 | 6.2% | 0.43 [0.11, 1.71] |
| Gante 2018 | 25 | 253 | 21 | 135 | 18.3% | 0.60 [0.32, 1.11] |
| Harper 2016 | 22 | 157 | 30 | 63 | 17.0% | 0.18 [0.09, 0.35] |
| Kahn 2006 | 10 | 77 | 5 | 18 | 7.5% | 0.39 [0.11, 1.32] |
| McGrath 2016 | 5 | 29 | 4 | 34 | 5.9% | 1.56 [0.38, 6.47] |
| Picón-César 2021 | 19 | 70 | 9 | 20 | 9.9% | 0.46 [0.16, 1.27] |
| Rowan 2008 | 38 | 195 | 56 | 168 | 23.0% | 0.48 [0.30, 0.78] |
| Yogev 2011 | 19 | 93 | 12 | 31 | 12.2% | 0.41 [0.17, 0.98] |
| Total (95% CI) | | 930 | | 482 | 100.0% | 0.43 [0.30, 0.63] |
| Total events | 147 | | 141 | | | |

Heterogeneity: $Tau^2 = 0.10$; $Chi^2 = 11.09$, $df = 7$ ($P = 0.13$); $I^2 = 37\%$
Test for overall effect: $Z = 4.36$ ($P < 0.0001$)

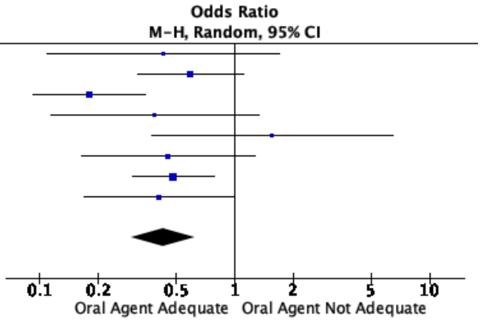


**Supplementary Figure 2.4B** Funnel Plot for Assessment of Publication Bias

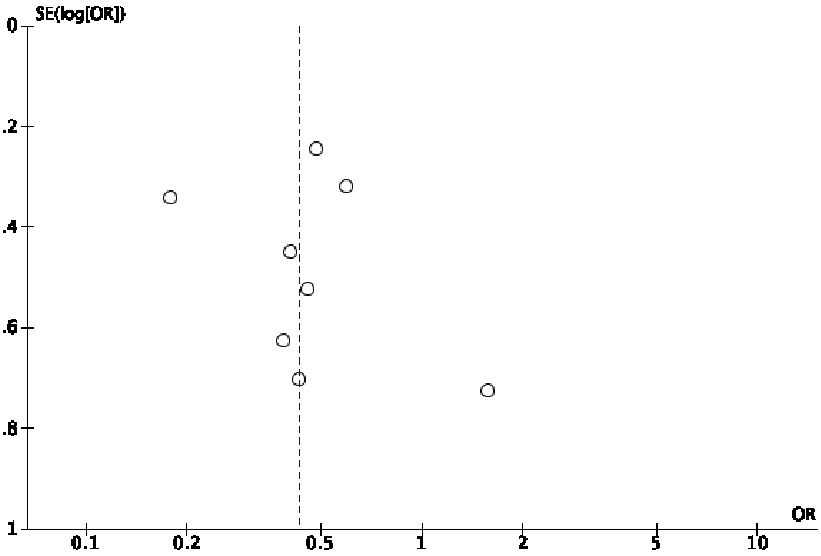

**Supplementary Figure 2.5A** Forest plot for included studies comparing if oral pharmacological agent was adequate or not adequate for haemoglobin A1C

| | Oral agent success | | | Oral agent failure | | | | Mean Difference |
|---|---|---|---|---|---|---|---|---|
| Study or Subgroup | Mean | SD | Total | Mean | SD | Total | Weight | IV, Random, 95% CI |
| Gante 2018 | 5.2 | 0.5 | 253 | 5.5 | 0.5 | 135 | 21.8% | −0.30 [−0.40, −0.20] |
| Khin 2018 | 5.4 | 0.33 | 61 | 5.6 | 0.48 | 77 | 17.3% | −0.20 [−0.34, −0.06] |
| McGrath 2016 | 5 | 0.3 | 29 | 5.2 | 0.4 | 34 | 13.1% | −0.20 [−0.37, −0.03] |
| Picón–César 2021 | 5.3 | 0.37 | 70 | 5.3 | 0.39 | 20 | 11.5% | 0.00 [−0.19, 0.19] |
| Rowan 2008 | 5.6 | 0.6 | 195 | 5.9 | 0.7 | 168 | 17.3% | −0.30 [−0.44, −0.16] |
| Tertti 2013 | 5.44 | 0.35 | 87 | 5.6 | 0.24 | 23 | 19.0% | −0.16 [−0.28, −0.04] |
| Total (95% CI) | | | 695 | | | 457 | 100.0% | −0.21 [−0.29, −0.13] |

Heterogeneity: $Tau^2 = 0.00$; $Chi^2 = 9.71$, $df = 5$ ($P = 0.08$); $I^2 = 49\%$
Test for overall effect: $Z = 5.17$ ($P < 0.00001$)

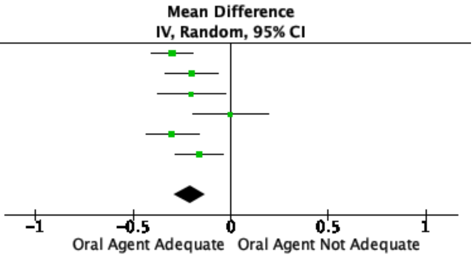


**Supplementary Figure 2.5B** Funnel Plot for Assessment of Publication Bias

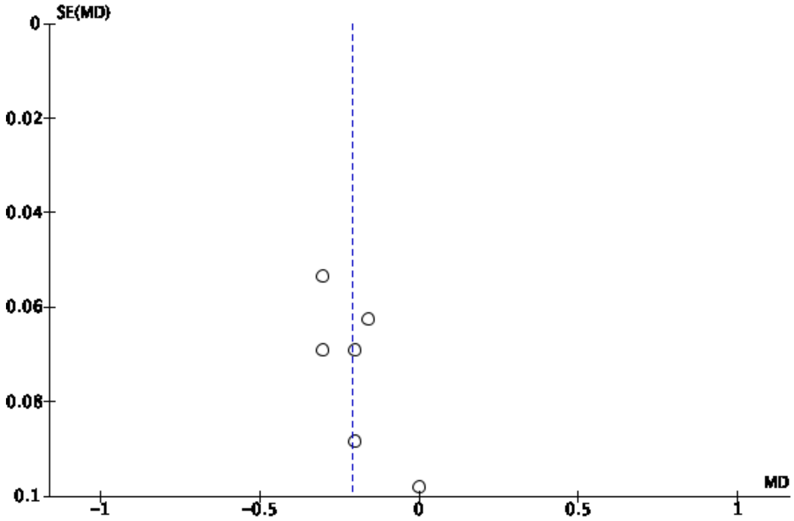

**Supplementary Figure 2.6A** Forest plot for included studies comparing if oral pharmacological agent was adequate or not adequate for fasting glucose

| | Oral agent success | | | Oral agent failure | | | | Mean Difference |
|---|---|---|---|---|---|---|---|---|
| Study or Subgroup | Mean | SD | Total | Mean | SD | Total | Weight | IV, Random, 95% CI |
| Chmait 2004 | 94 | 15 | 56 | 105 | 22 | 13 | 5.3% | −11.00 [−23.59, 1.59] |
| Conway 2004 | 102 | 14 | 63 | 115 | 24 | 12 | 4.7% | −13.00 [−27.01, 1.01] |
| Gante 2018 | 85.4 | 11.41 | 253 | 92.7 | 15.3 | 135 | 11.2% | −7.30 [−10.24, −4.36] |
| Harper 2016 | 106 | 15 | 157 | 138 | 52 | 63 | 5.1% | −32.00 [−45.05, −18.95] |
| Kahn 2006 | 100 | 17 | 77 | 112 | 24 | 18 | 5.7% | −12.00 [−23.72, −0.28] |
| Khin 2018 | 86.9 | 11.9 | 61 | 97.2 | 16.6 | 77 | 10.2% | −10.30 [−15.06, −5.54] |
| McGrath 2016 | 86.4 | 10.8 | 29 | 88.2 | 10.8 | 34 | 9.8% | −1.80 [−7.15, 3.55] |
| Picón-César 2021 | 91.71 | 10.81 | 70 | 97.48 | 9.91 | 20 | 10.0% | −5.77 [−10.80, −0.74] |
| Rochon 2006 | 102 | 22 | 80 | 107 | 15 | 21 | 7.9% | −5.00 [−13.02, 3.02] |
| Rowan 2008 | 95.4 | 14.4 | 195 | 109.8 | 19.8 | 168 | 10.9% | −14.40 [−18.01, −10.79] |
| Tertti 2013 | 99 | 9 | 87 | 102.6 | 12.6 | 23 | 9.7% | −3.60 [−9.09, 1.89] |
| Yogev 2011 | 95.9 | 16.2 | 93 | 91.7 | 12.6 | 31 | 9.7% | 4.20 [−1.32, 9.72] |
| Total (95% CI) | | | 1221 | | | 615 | 100.0% | −8.02 [−11.87, −4.16] |

Heterogeneity: $Tau^2 = 32.19$; $Chi^2 = 54.70$, df = 11 ($P < 0.00001$); $I^2 = 80\%$
Test for overall effect: Z = 4.07 ($P < 0.0001$)

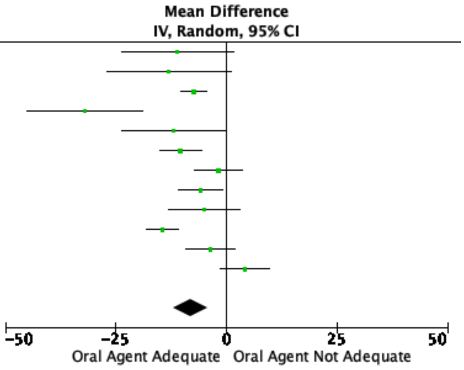


**Supplementary Figure 2.6B** Funnel Plot for Assessment of Publication Bias

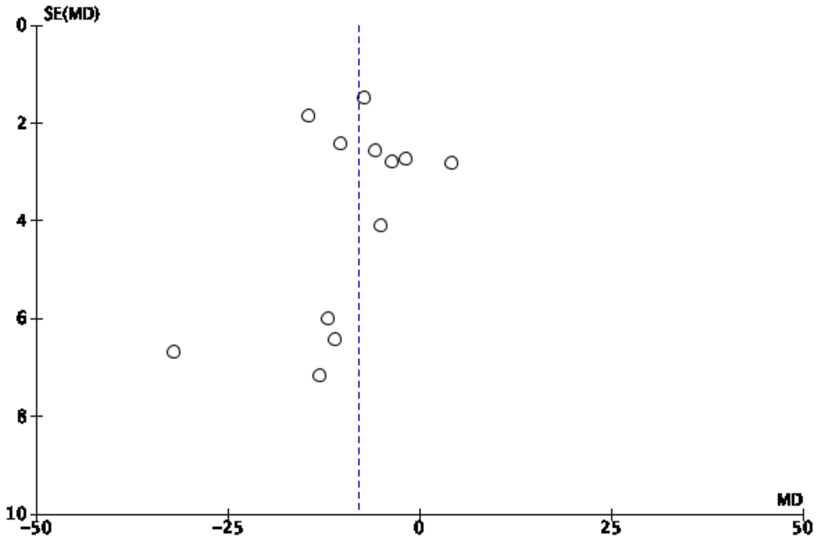

**Supplementary Figure 2.7A** Forest plot for included studies comparing if oral pharmacological agent was adequate or not adequate for 1-hour glucose

| | Oral agent success | | | Oral agent failure | | | | Mean Difference |
|---|---|---|---|---|---|---|---|---|
| Study or Subgroup | Mean | SD | Total | Mean | SD | Total | Weight | IV, Random, 95% CI |
| Chmait 2004 | 199 | 25 | 56 | 206 | 58 | 13 | 4.4% | −7.00 [−39.20, 25.20] |
| Conway 2004 | 205 | 23 | 63 | 230 | 36 | 12 | 7.9% | −25.00 [−46.15, −3.85] |
| Gante 2018 | 171.4 | 32.2 | 253 | 176.8 | 33.1 | 135 | 17.3% | −5.40 [−12.25, 1.45] |
| Harper 2016 | 201 | 28 | 157 | 218 | 31 | 63 | 15.9% | −17.00 [−25.82, −8.18] |
| Picón-César 2021 | 198.92 | 30.81 | 70 | 217.31 | 9.64 | 20 | 16.2% | −18.39 [−26.75, −10.03] |
| Rochon 2006 | 200 | 33 | 80 | 223 | 31 | 21 | 11.2% | −23.00 [−38.10, −7.90] |
| Tertti 2013 | 201.6 | 25.2 | 87 | 201.6 | 28.8 | 23 | 12.8% | 0.00 [−12.91, 12.91] |
| Yogev 2011 | 198.8 | 29.4 | 93 | 193 | 25.3 | 31 | 14.4% | 5.80 [−4.92, 16.52] |
| Total (95% CI) | | | 859 | | | 318 | 100.0% | −10.64 [−18.25, −3.02] |

Heterogeneity: $Tau^2 = 75.01$; $Chi^2 = 23.47$, df = 7 ($P = 0.001$); $I^2 = 70\%$

Test for overall effect: Z = 2.74 ($P = 0.006$)

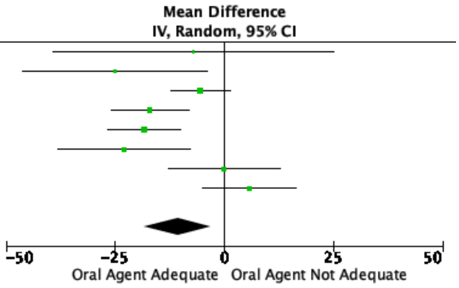


**Supplementary Figure 2.7B** Funnel Plot for Assessment of Publication Bias

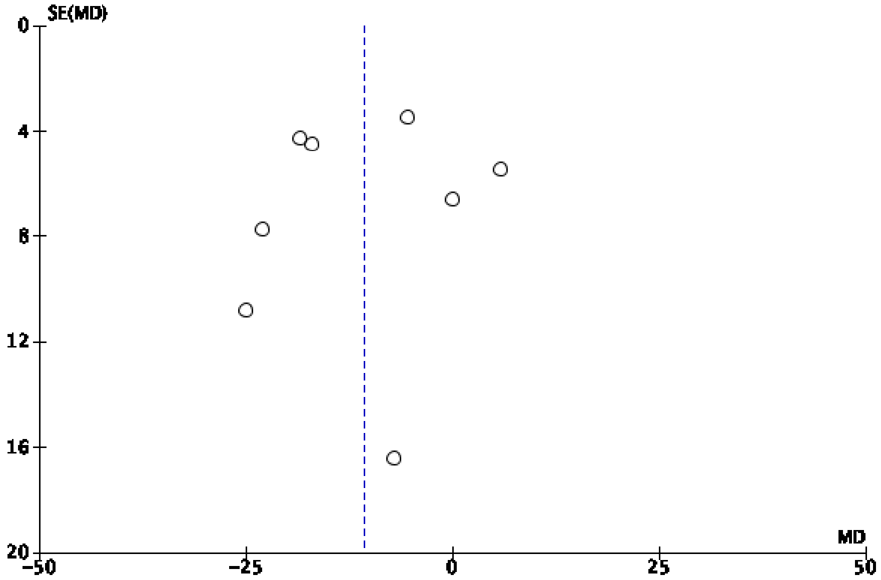

**Supplementary Figure 2.8A** Forest plot for included studies comparing if oral pharmacological agent was adequate or not adequate for 2-hour glucose

| | Oral agent success | | | Oral agent failure | | | | Mean Difference |
|---|---|---|---|---|---|---|---|---|
| Study or Subgroup | Mean | SD | Total | Mean | SD | Total | Weight | IV, Random, 95% CI |
| Chmait 2004 | 169 | 31 | 56 | 192 | 25 | 13 | 6.2% | −23.00 [−38.83, −7.17] |
| Conway 2004 | 169 | 34 | 63 | 204 | 66 | 12 | 1.1% | −35.00 [−73.27, 3.27] |
| Gante 2018 | 144.6 | 33.8 | 253 | 152.9 | 40 | 135 | 21.2% | −8.30 [−16.23, −0.37] |
| Harper 2016 | 178 | 28 | 157 | 188 | 42 | 63 | 11.7% | −10.00 [−21.26, 1.26] |
| Khin 2018 | 154.6 | 21.2 | 61 | 159.7 | 27 | 77 | 20.8% | −5.10 [−13.14, 2.94] |
| McGrath 2016 | 153 | 25.2 | 29 | 149.4 | 25.2 | 34 | 9.7% | 3.60 [−8.88, 16.08] |
| Picón–César 2021 | 185.41 | 25.77 | 70 | 195.14 | 32.25 | 20 | 6.6% | −9.73 [−25.10, 5.64] |
| Rochon 2006 | 179 | 33 | 80 | 189 | 46 | 21 | 3.6% | −10.00 [−30.96, 10.96] |
| Tertti 2013 | 147.6 | 32.4 | 87 | 153 | 28.8 | 23 | 8.3% | −5.40 [−19.00, 8.20] |
| Yogev 2011 | 165.9 | 31.8 | 93 | 169.6 | 27.8 | 31 | 10.8% | −3.70 [−15.43, 8.03] |
| Total (95% CI) | | | 949 | | | 429 | 100.0% | −7.31 [−11.38, −3.25] |

Heterogeneity: $Tau^2 = 3.88$; $Chi^2 = 9.88$, df = 9 ($P = 0.36$); $I^2 = 9\%$
Test for overall effect: $Z = 3.53$ ($P = 0.0004$)

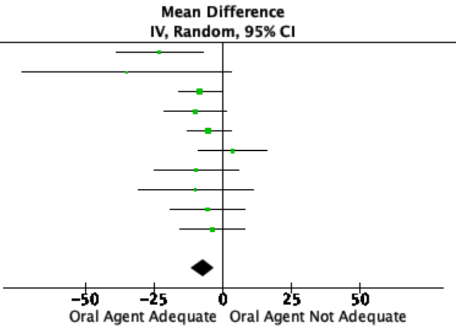


**Supplementary Figure 2.8B** Funnel Plot for Assessment of Publication Bias

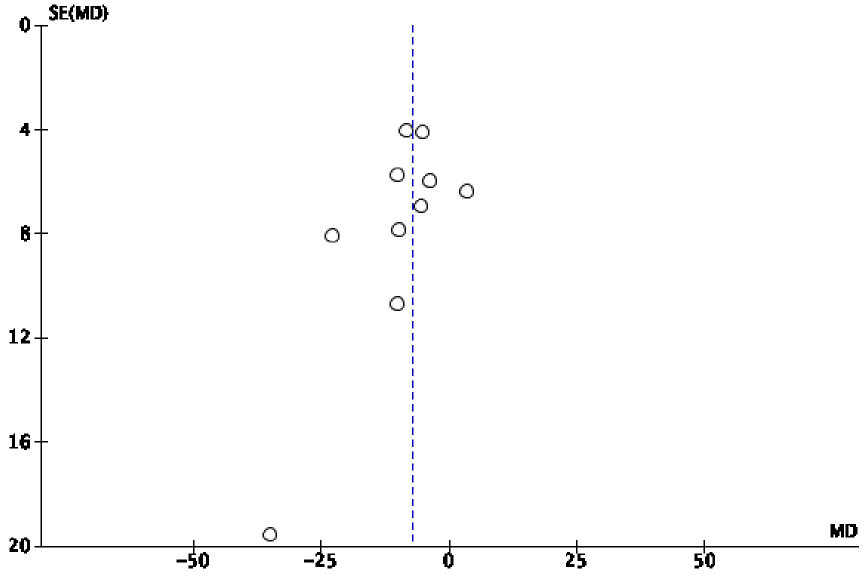

**Supplementary Figure 2.9A** Forest plot for included studies comparing if oral pharmacological agent was adequate or not adequate for 3-hour glucose

| Study or Subgroup | Oral agent success Mean | SD | Total | Oral agent failure Mean | SD | Total | Weight | Mean Difference IV, Random, 95% CI |
|---|---|---|---|---|---|---|---|---|
| Chmait 2004 | 126 | 33 | 56 | 128 | 45 | 13 | 12.3% | −2.00 [−27.94, 23.94] |
| Conway 2004 | 133 | 33 | 63 | 176 | 65 | 12 | 7.4% | −43.00 [−80.67, −5.33] |
| Harper 2016 | 140 | 27 | 157 | 143 | 42 | 63 | 23.9% | −3.00 [−14.20, 8.20] |
| Picón-César 2021 | 150.81 | 29.37 | 70 | 147.21 | 43.96 | 20 | 15.9% | 3.60 [−16.86, 24.06] |
| Rochon 2006 | 138 | 35 | 80 | 114 | 38 | 21 | 17.8% | 24.00 [6.03, 41.97] |
| Yogev 2011 | 118 | 36.3 | 93 | 121.1 | 28.9 | 31 | 22.6% | −3.10 [−15.67, 9.47] |
| Total (95% CI) | | | 519 | | | 160 | 100.0% | 0.00 [−11.79, 11.79] |

Heterogeneity: $Tau^2 = 118.73$; $Chi^2 = 12.50$, $df = 5$ ($P = 0.03$); $I^2 = 60\%$
Test for overall effect: $Z = 0.00$ ($P = 1.00$)

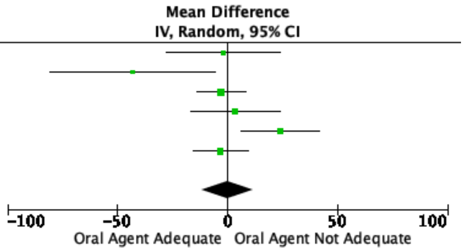


**Supplementary Figure 2.9B** Funnel Plot for Assessment of Publication Bias

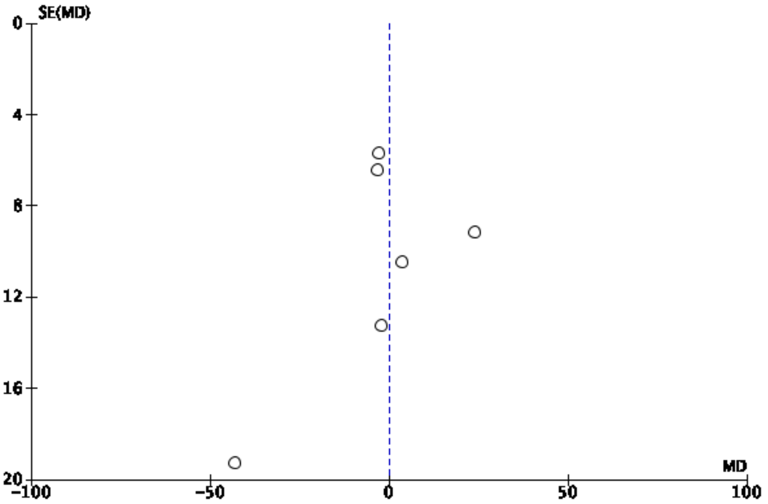

**Supplementary Figure 2.10A** Forest plot for included studies comparing if oral pharmacological agent was adequate or not adequate for family history of diabetes

| Study or Subgroup | Oral agent success Events | Oral agent success Total | Oral agent failure Events | Oral agent failure Total | Weight | Odds Ratio M-H, Random, 95% CI |
|---|---|---|---|---|---|---|
| Chmait 2004 | 23 | 50 | 5 | 11 | 9.4% | 1.02 [0.28, 3.79] |
| Conway 2004 | 46 | 63 | 10 | 12 | 6.7% | 0.54 [0.11, 2.73] |
| Gante 2018 | 114 | 253 | 53 | 135 | 29.0% | 1.27 [0.83, 1.94] |
| McGrath 2016 | 14 | 29 | 15 | 34 | 13.8% | 1.18 [0.44, 3.20] |
| Picón-César 2021 | 37 | 70 | 15 | 20 | 11.8% | 0.37 [0.12, 1.14] |
| Rowan 2008 | 74 | 195 | 88 | 168 | 29.3% | 0.56 [0.37, 0.85] |
| Total (95% CI) | | 660 | | 380 | 100.0% | 0.79 [0.50, 1.25] |
| Total events | 308 | | 186 | | | |

Heterogeneity: $Tau^2 = 0.14$; $Chi^2 = 10.15$, $df = 5$ ($P = 0.07$); $I^2 = 51\%$
Test for overall effect: $Z = 1.00$ ($P = 0.32$)

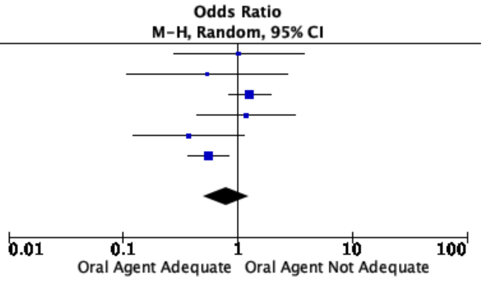


**Supplementary Figure 2.10B** Funnel Plot for Assessment of Publication Bias

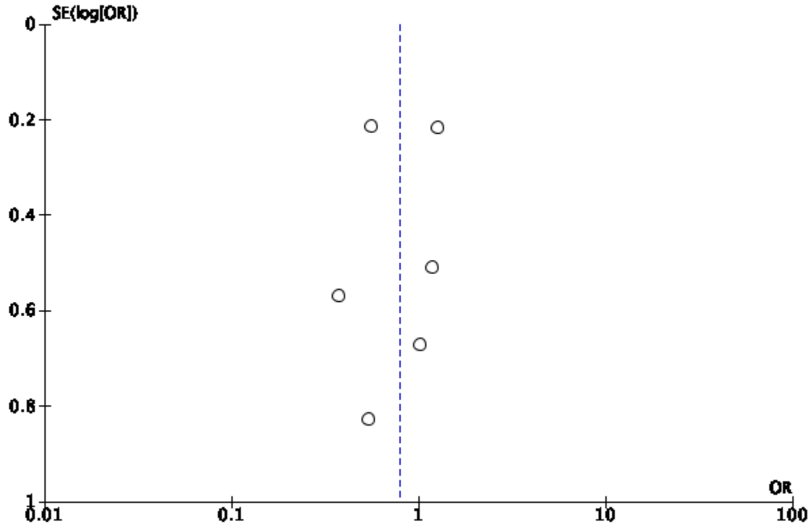

**Supplementary Figure 2.11A** Forest plot for included studies comparing if oral pharmacological agent was adequate or not adequate for gestational age at gestational diabetes diagnosis

| Study or Subgroup | Oral agent success Mean | SD | Total | Oral agent failure Mean | SD | Total | Weight | Mean Difference IV, Random, 95% CI |
|---|---|---|---|---|---|---|---|---|
| Chmait 2004 | 27.3 | 7.5 | 56 | 20 | 8.6 | 13 | 4.3% | 7.30 [2.23, 12.37] |
| Conway 2004 | 20 | 8.3 | 63 | 18.4 | 8.4 | 12 | 4.2% | 1.60 [−3.58, 6.78] |
| Gante 2018 | 20.3 | 8.1 | 253 | 17.5 | 8 | 135 | 12.6% | 2.80 [1.12, 4.48] |
| Harper 2016 | 25.7 | 3.7 | 157 | 19.6 | 7.8 | 63 | 11.4% | 6.10 [4.09, 8.11] |
| Kahn 2006 | 28 | 5 | 77 | 22.7 | 7 | 18 | 7.2% | 5.30 [1.88, 8.72] |
| Khin 2018 | 27 | 5 | 61 | 25 | 6 | 77 | 12.0% | 2.00 [0.16, 3.84] |
| McGrath 2016 | 24 | 6 | 29 | 23 | 6 | 34 | 8.3% | 1.00 [−1.97, 3.97] |
| Picón-César 2021 | 23.48 | 6.15 | 70 | 22.19 | 6.82 | 20 | 7.4% | 1.29 [−2.03, 4.61] |
| Rochon 2006 | 26 | 7 | 80 | 24 | 7 | 21 | 7.3% | 2.00 [−1.36, 5.36] |
| Tertti 2013 | 27.1 | 2.3 | 87 | 25.7 | 3.2 | 23 | 13.6% | 1.40 [0.01, 2.79] |
| Yogev 2011 | 26.5 | 2.8 | 93 | 25.9 | 5.3 | 31 | 11.6% | 0.60 [−1.35, 2.55] |
| Total (95% CI) | | | 1026 | | | 447 | 100.0% | 2.64 [1.42, 3.86] |

Heterogeneity: $Tau^2 = 2.34$; $Chi^2 = 26.42$, df = 10 (P = 0.003); $I^2 = 62\%$
Test for overall effect: Z = 4.24 (P < 0.0001)

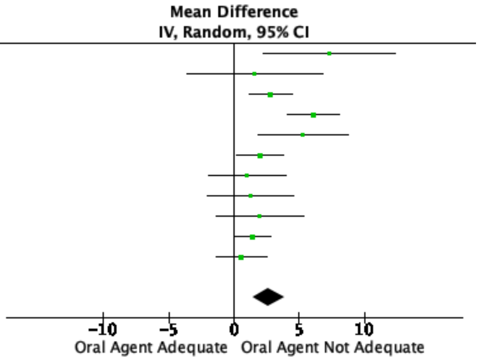


**Supplementary Figure 2.11B** Funnel Plot for Assessment of Publication Bias

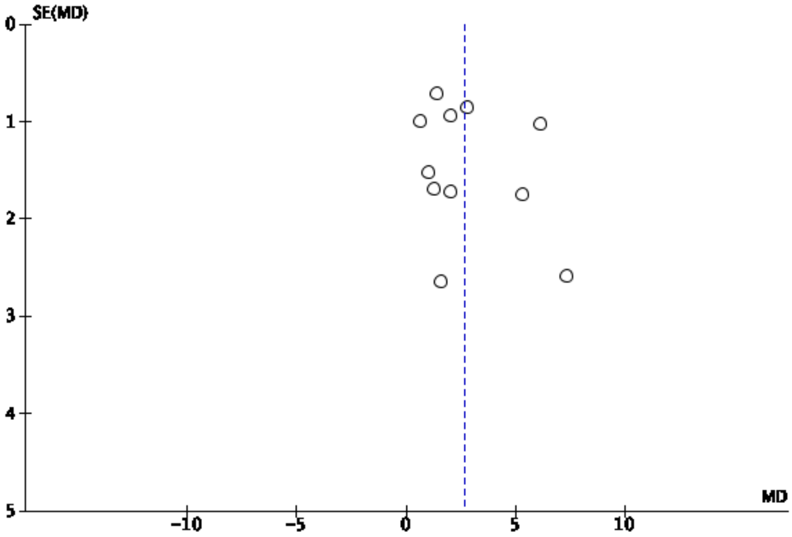

**Table 2.12B** Forest plot for included studies comparing if oral pharmacological agent was adequate or not adequate for gestational age at initiation of oral pharmacological agent for treatment of GDM

| Study or Subgroup | Oral agent success Mean | SD | Total | Oral agent failure Mean | SD | Total | Weight | Mean Difference IV, Random, 95% CI |
|---|---|---|---|---|---|---|---|---|
| Chmait 2004 | 29.7 | 6.6 | 56 | 23 | 6.9 | 13 | 9.4% | 6.70 [2.57, 10.83] |
| Gante 2018 | 28.8 | 6.1 | 253 | 24.3 | 6.6 | 135 | 18.2% | 4.50 [3.16, 5.84] |
| Kahn 2006 | 31 | 4 | 77 | 24 | 7 | 18 | 11.5% | 7.00 [3.65, 10.35] |
| Khin 2018 | 29 | 4 | 61 | 26 | 6 | 77 | 17.1% | 3.00 [1.33, 4.67] |
| McGrath 2016 | 28.9 | 4.9 | 29 | 25.3 | 5.8 | 34 | 13.8% | 3.60 [0.96, 6.24] |
| Picón-César 2021 | 28.23 | 5.01 | 70 | 24.6 | 6.92 | 20 | 11.8% | 3.63 [0.38, 6.88] |
| Yogev 2011 | 30.1 | 2.7 | 93 | 29.5 | 3.3 | 31 | 18.3% | 0.60 [-0.68, 1.88] |
| Total (95% CI) | | | 639 | | | 328 | 100.0% | 3.79 [2.08, 5.51] |

Heterogeneity: $Tau^2 = 3.76$; $Chi^2 = 27.06$, df = 6 (P = 0.0001); $I^2 = 78\%$
Test for overall effect: Z = 4.33 (P < 0.0001)

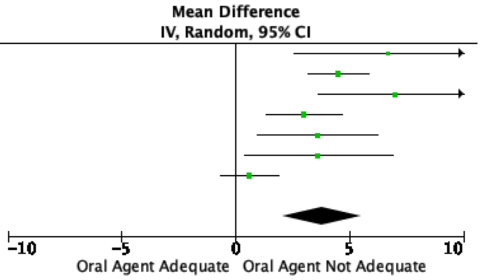


**Supplementary Figure 2.12B** Funnel Plot for Assessment of Publication Bias

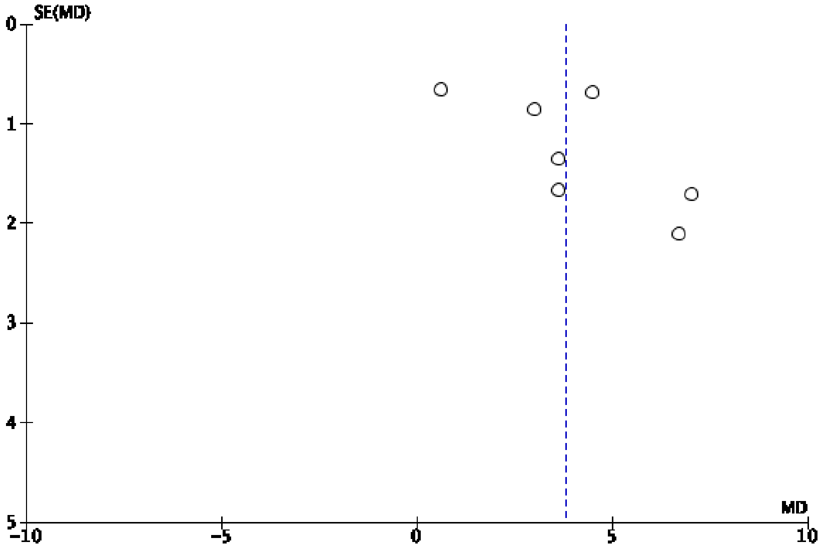

**Supplementary Table 1 Narrative summary of studies not included in the meta-analysis**

| Author | Key findings |
|---|---|
| *Insulin required when diet not adequate* | |
| Berg 2007[24] | The need for insulin treatment increased with increasing BMI. Among women with normal weight (BMI <25 kg/m$^2$), 9.6% (n=33) required insulin, compared to 31.9% (n=52) in the obese group (p <0.001) |
| Elnour 2008[27] | The number of abnormal OGTT values, specifically fasting values (≥95 mg/dl) and 1-h values (≥180 mg/dl), contributed significantly to insulin need during the index pregnancy (P<0.05). |
| Giannubilo 2018[28] | The overall risk of need for insulin therapy was significantly higher in women carrying a male fetus (odds ratio = 1.837; 95% CI, 1.737–2.8775; P = 0.0078) |
| Gibson 2012[29] | There were no significant differences in age, race, or parity among patients requiring insulin therapy and diet/exercise-controlled GDM. Patients requiring insulin therapy had significantly higher pre-pregnancy BMI, but comparable 1-h glucose tolerance test, gestational age at delivery, and weight gain. |
| Hillier 2013[3030] | Treatment with insulin more likely in higher BMI groups and non-white ethnicities |
| Molina-Vega 2020[3737] | Greater BMI was associated with greater odds of needing insulin therapy (OR 1.103; p<0.001). Age (OR 1.019; p=0.429) and pregnancy during winter (OR 0.493; p=0.050, spring (OR 0.626; p=0.159) or summer (0.680; p=0.214), as opposed to during fall were not associated with the need for insulin therapy. |
| Nguyen 2016[3939] | Maternal predictors of antepartum insulin therapy:<br>Pre-pregnancy BMI (OR 1.03; 95% CI 1.01 – 1.06), Past history of GDM (OR 1.77; 9%% CI 1.24–2.56), Diagnosis of GDM <20 weeks gestation (OR 3.32; 95% CI 1.87–6.19), fasting plasma glucose (OR 1.65; 95% CI 1.10–2.47), 2-hour post-OGTT glucose (OR 1.39; 95% CI 0.91–2.14); Both glucose values above local reference range at screening (OR 2.60; 95% CI 1.98 – 3.44)<br>Maternal factors not significantly associated with insulin therapy:<br>Past history of caesarean delivery (OR 1.41; 95% CI 0.96 – 2.08), Past history of macrosomia (OR 0.79; 95%CI 0.88 – 1.46) |
| Parrettini 2020[42] | Maternal age and distribution of diagnostic values at the OGTT were the only factors that were significantly associated with a risk of need for insulin therapy (p < 0.05). |
| Wong 2012[47] | Women from South-East Asia had the lowest need to start insulin (37.2%), compared with Anglo-Europeans (56.7%, P < 0.001) - SE Asian (37.2%), South Asian (55%), Middle Eastern (51.6%), Anglo-European (56.7%), Pacific Islander (65.5%) - ANOVA p<0.001<br>Women from South-East Asia had the lowest need for rapid-acting insulin for the management of postprandial hyperglycaemia (30.4% vs. 44.9% for women from Anglo-European background, P = 0.002). Pacific Islanders had the greatest need for insulin therapy, but were started on insulin at a later stage. |
| Zawiejska 2014[49] | Maternal fasting hyperglycaemia >=5.1 mmol/l increased odds of requiring insulin therapy (OR 3.8 [95% CI 2.3, 6.5]; aOR 2.6 [1.4, 4.9]) |
| *Pharmacological therapy required (oral agent and/or insulin) when diet not adequate* | |

| | |
|---|---|
| Zhu 2021[54] | The strongest predictor of moving from diet to medication was the 2nd trimester FBG, recording an odds ratio of 3.58. This suggests that for every additional 1 mmol/L the FBG was elevated, patients were 3.58 times more likely be medicated for their GDM, controlling for other factors in the model. The other significant predictor was age, with an odds ratio of 1.06. |
| *Genetic predictors of glyburide being inadequate to achieve target glucose values* | |
| Bouchghoul et al 2021[67] | CYP2C9 and OATP1B3 genetic polymorphisms<br>The percentage of patients who switched from glyburide to insulin was much higher (× 1.8) in the variant genotype group, although this was not statistically significant: 23.8% (5/21) vs. 13.0% (7/54) in the wild-type genotype group and 19.0% (8/42) in the intermediate group (trend test-logistic regression, P = 0.24).<br>The fasting glycaemic control of diabetes with glyburide was, on average, better in the wild-type genotype group than in the intermediate group and the variant group, with a lower percentage of out-of-target blood glucose values |
| *Maternal lipidome responses to metformin and insulin treatment* | |
| Huhtala et al 2020[68] | Fasting serum lipidome measured at diagnosis (mean 30 weeks gestation) and at 36 weeks using nuclear magnetic resonance spectroscopy<br>Compared to insulin, metformin treatment of GDM led to higher maternal serum concentrations of triglyceride-rich lipoproteins. Especially triglycerides and cholesterol in VLDL were positively associated with birthweight.<br>Women with high VLDL cholesterol or high apoB/apoA-1 may benefit from insulin treatment over metformin with respect to offspring birthweight |
